# User and Usability Testing of a web-based *GBA* and *LRRK2* genetics education tool

**DOI:** 10.1101/2022.12.09.22283291

**Authors:** Noah Han, Rachel A. Paul, Tanya Bardakjian, Daniel Kargilis, Angela R. Bradbury, Alice Chen-Plotkin, Thomas F. Tropea

## Abstract

**Background:** Genetic testing is essential to identify research participants for clinical trials enrolling people with Parkinson’s disease (PD) carrying a variant in the *GBA* or *LRRK2* genes. Limited availability of professionals trained in genetic counseling is a major barrier to increased testing.

**Objective:** As an alternative to pre-test genetic counseling, we developed a web-based genetics education tool focused on *GBA* and *LRRK2* testing for PD called the Interactive Multimedia Approach to Genetic counseling to INform and Educate in PD (IMAGINE-PD) and conducted user testing and usability testing.

**Methods:** Genetic counselors and PD and neurogenetics subject matter experts developed content for IMAGINE-PD, specifically focused on *GBA* and *LRRK2* genetic testing. User testing to review content was conducted with thirteen patients with PD and eleven movement disorder specialists (MDS). Twelve PD patients were enrolled in usability testing of a high-fidelity prototype in accordance with national guidelines. Qualitative data analysis was used in both phases.

**Results:** Qualitative analysis conducted by three evaluators identified themes emerging from MDS and PD patient feedback in user testing in three content areas: content, function, and appearance. Similarly, qualitative analysis of usability testing feedback identified additional themes in these three content areas. Refinements were made to IMAGINE-PD based on consensus recommendation by evaluators.

**Conclusions:** User testing for content review and usability testing have informed refinements to IMAGINE-PD to develop this alternative to standard pre-test genetic counseling. Comparison of this stakeholder-informed intervention to standard telegenetic counseling approaches is ongoing.

## Introduction

Parkinson’s Disease (PD) is the second commonest neurodegenerative disease and the fastest growing neurological disease worldwide.[1, 2] Variants in *LRRK2*, *GBA*, *Parkin*, *DJ-1*, *VPS-35, Pink1,* and *SNCA* are identified in 10-12% of PD cases.[3–9] Despite a genetic mutation frequency similar to some cancer syndromes where germline genetic testing is common,[10, 11] genetic testing is not standard in the evaluation and management of PD and is rarely conducted as part of clinical care.[12] However, knowing one’s genetic status is already of key importance for research, as therapies targeting carriers of *GBA* and *LRRK2* variants are in clinical trials.[13, 14] Additionally, PD patients have expressed interest in learning their genetic information.[15, 16]

Recently proposed recommendations would expand clinical or research genetic testing to nearly all PD patients.[17] However, the standard service delivery model for genetic testing includes pre-test and post-test genetic counseling, which is limited by the availability of specialized genetic counselors or physicians with sufficient genetics training.[18] Indeed, there are only 125 genetic counselors specialized in neurogenetics offering in-person visits, and 82 genetic counselors offering telehealth visits listed on the National Society of Genetic Counselors (NSGC)’s public directory for genetic counselors (findageneticcounselor.nsgc.org). We developed a web-based education tool focused on *GBA* and *LRRK2* genetic testing called the Interactive, Multimedia Approach to Genetic counseling to INform and Educate in Parkinson’s Disease (IMAGINE-PD) to address this gap. IMAGINE-PD could be made widely available to increase genetics education prior to *GBA* and *LRRK2* testing in clinical or research settings to identify research-eligible PD patients.

The goal of this work is to create a genetics education tool for *GBA* and *LRRK2* testing incorporating key stakeholder input. Structured interviews were conducted with movement disorder specialists (MDS) and PD patients to evaluate the content of IMAGINE-PD in user testing, and with PD patients to evaluate website usability according to the U.S. Department of Health and Human Services (DHHS) Research-Based Web Design & Usability Guidelines guidelines.[19] Qualitative data analysis informed changes to create a final version of IMAGINE-PD.

## Materials and Methods

### IMAGINE-PD Web Development

Website development and usability testing were conducted in accordance with the DHHS research-based web design and usability guidelines.[19] Core content for a genetic counseling tool for *GBA* and *LRRK2* variant genetic testing for PD was developed by the authors (T.B., A.C.P, and T.F.T). *GBA* and *LRRK2* variant testing information was included to align with the research-based testing being performed as part of the MIND Initiative research study ongoing at UPenn.[20] Content was then assigned to ‘primary’ or essential, ‘secondary’ or important but not essential, or ‘optional’ categories based on their level of importance. Primary concepts included a review of PD, basic genetics concepts, genetics of PD, genetic results disclosure, and limitations and implications of genetic testing. For each primary concept, an audiovisual recording of a movement disorders physician or genetic counselor describing the concept was created. Secondary information included: diagnosis, symptoms, and treatment of PD, an introduction to genetic counseling, a review of genetic testing (GT), types of genetic tests and results, risks and benefits of GT, a review of *GBA* including Gaucher’s disease and *LRRK2*, and a review of other genetic causes of PD that are not tested in the study (limitations). One or more slides were created to capture the secondary content important for genetic counseling including text, visuals, and an optional audio recording of the text presented. Links to outside reading were identified for optional material. All content was organized into a high-fidelity prototype website using http://Wix.com (Tel Aviv, Israel). Once the preliminary set of material was created, we solicited feedback on content and website organization from genetic counselors and physician-scientists with expertise in neurogenetics. Changes were made to address points of feedback. Working with the UPenn Center for Clinical Epidemiology and Biostatistics Clinical Research Computing Unit, we created a new website incorporating the content from the prototype compatible with common web-browsers and operating systems, accessible on typical connection speeds, across a range of different resolutions and orientations, to account for use on computer, smart phone, or tablet computer. Number of visits and time spent on each page is captured. A computer graphics specialist with experience in medical artwork created animations (http://spmotiondesign.com). Readability of each page was evaluated using readable.com to ensure a Flesch-Kincaid readability index of below 9. With this prototype, user testing was first conducted, and changes were then incorporated into IMAGINE-PD after reviewing feedback. Usability testing was conducted subsequently, and again feedback was reviewed and refinements were made to the content, organization, and web interface. The final version is available at http://www.pennmedicine.org/imaginepd. A flowchart of the IMAGINE-PD web development and testing process can be viewed in **Figure 1**.

**Figure 1.**
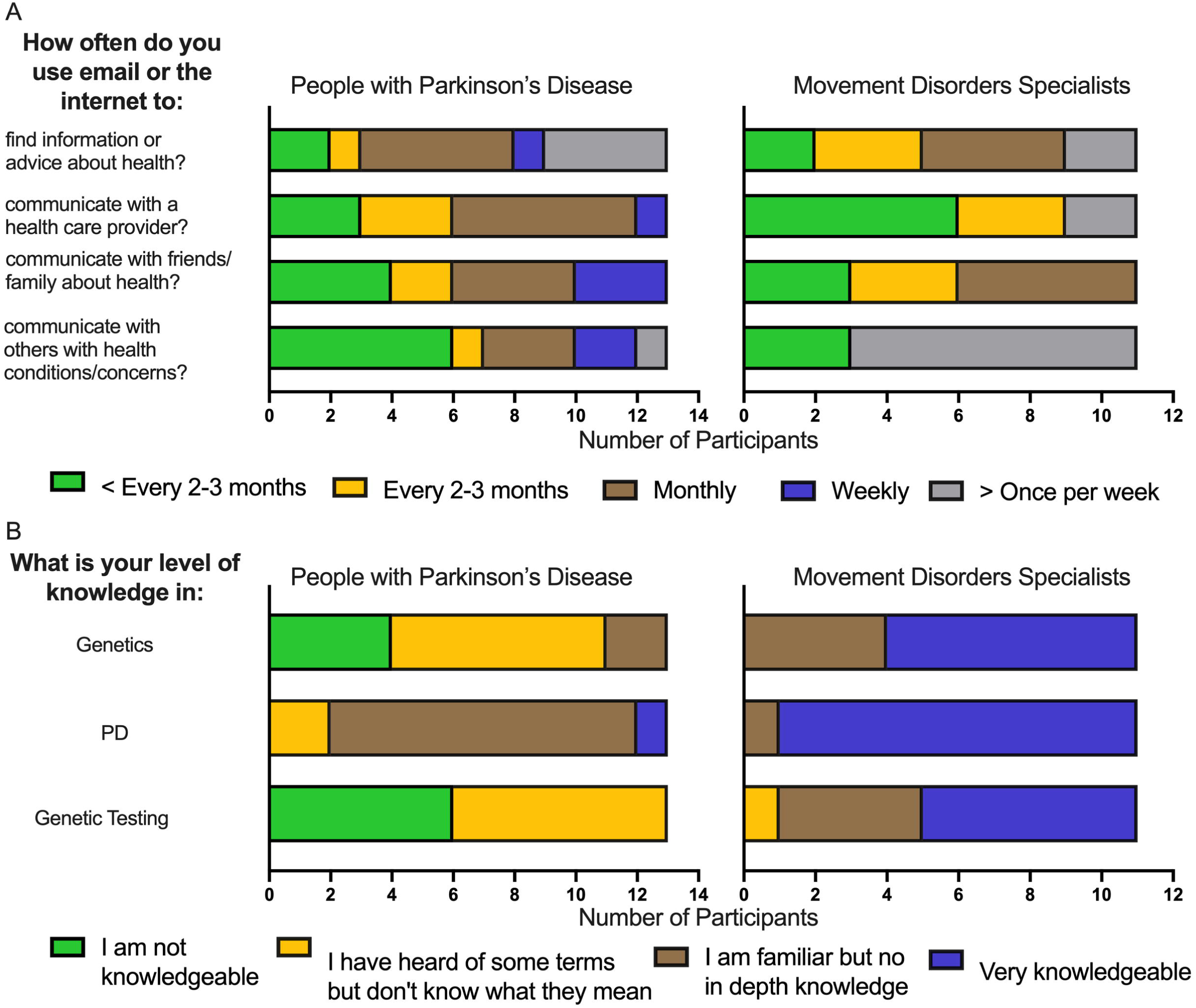
IMAGINE-PD Website development flowchart.

### Participants

In the user testing phase, thirteen cognitively normal patients with PD who receive care at UPenn, referred by their physicians, were enrolled. Movement disorders specialists (MDS) at the UPenn Parkinson’s Disease and Movement Disorders Center (PDMDC) and the Philadelphia Veterans Affairs Parkinson’s Disease Research, Education, and Clinical Center (PADRECC) were asked to participate (excluding authors ACP and TFT) for a total of eleven neurologists or psychiatrists. In the usability testing phase, twelve PD patients who did not participate in the user testing phase were enrolled. In both phases, sample sizes exceeded the suggestions outlined in the DHHS usability testing guidelines.[19] Identified participants were approached in person, by phone, or by email. Informed consent was obtained, and study visits were performed in-person by a single evaluator (NH).

### User Testing

Structured interviews were conducted between a single evaluator with experience in conducting clinical research visits (NH) and a participant between 10/31/2019 and 12/11/2019. The evaluator had no prior relationship with any participant, nor any assumptions or presuppositions about the outcomes of the interviews. First, participants were asked eleven questions about their general internet use, four questions about e-mail and internet use for health information (adapted from Baker et al),[21] and three questions assessing background knowledge in genetics, PD, and genetic testing. During the interview the evaluator asked questions of each participant, and data were entered data directly into RedCap.[22, 23] The evaluator navigated to each webpage in the prototype allowing the participant to view and listen to all material on that page with unlimited time. Prompts were given to ensure all participants interacted with all aspects of each page. Feedback was solicited using four open-ended questions, a 1-10 rating of usefulness, and a 1-4 rating of clarity of presentation. After all webpages were reviewed, seven additional questions were asked pertaining to the entire series of webpages and to solicit overall comments. Questionnaires are available in the **Supplement**.

### Usability Testing

The second iteration of IMAGINE-PD was created after the results of user testing was reviewed, and changes were implemented. For usability testing, structured videoconference interviews (https://bluejeans.com) were conducted between evaluator (NH) and participant between 8/18/2020 and 9/14/2020. Again, the evaluator had no prior relationship with any participant, nor any assumptions or presuppositions about the outcomes of the interviews. First, participants were asked an abbreviated eight question internet use questionnaire. The participant navigated to each webpage with unlimited time to view all content. Participants were asked to use the ‘share-screen’ function so the evaluator could observe their navigation of the website.

Prompts were given to ensure all participants interacted with all aspects of each page. Feedback was solicited using eight open-ended questions asked for each page. Questionnaires are available in the **Supplement**.

### Data Analysis

Descriptive statistics are presented for all demographic data and scales. A qualitative analysis plan based on a grounded theory was developed in advance of data review in consultation with Judy Shea, PhD. For both user testing review and usability testing phases all data were collected into a spreadsheet excluding identifying information. Three evaluators (NH, RAP, TFT) were given the following rules: 1) review the data from each webpage for PD providers and PD patients separately and identify 1 or more key themes per page, if a theme is apparent, and 2) if more than 1 theme is apparent, order themes for each page in order of importance or frequency. Data was independently reviewed by each reviewer to improve trustworthiness of the analysis. Subsequently, the three evaluators met to establish consensus on the key themes. Suggestions for changes to the website were made after all themes were reviewed and discussed among the evaluators.

### Standard Protocol Approvals, Registrations, and Patient Consents

Institutional Review Board (IRB) approval was obtained before initiating the study and informed consent was obtained from all participants prior to any study activities. This research adheres to the principles set out in the Declaration of Helsinki.

### Data Availability

All deidentified data can be made available upon request of the authors.

## Results

### Website Development

The final version includes 27 webpages, including three video-only pages and three video pages with animations. Audiovisual pages also include accessible text for participants with hearing impairment. Each non-video page included audio recording of the text for auditory learners. The website is accessible via computer browser, mobile device, or tablet. The length of time to view all videos is 7 minutes and 42 seconds and estimated time to review all website content is 25 – 40 min. The Flesch-Kincaid reading level for all pages was median of 8.3 (7.375– 8.25).

### User Testing

Participant demographics are summarized in **Table 1**. E-mail and internet use for health information is described in **Figure 2A** and **Supplemental Table 1** in the Supplement. PD, genetics, and genetic testing knowledge is reported in **Figure 2B**.

**Table 1.**
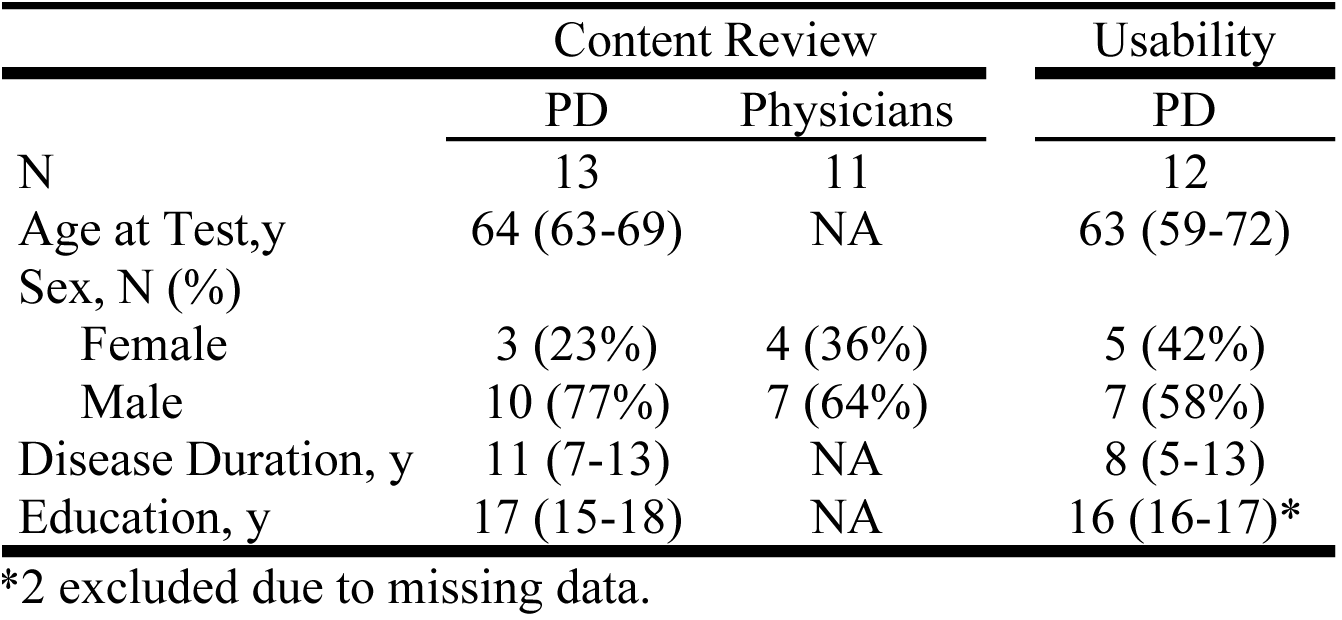
Cohort Description

**Figure 2.**
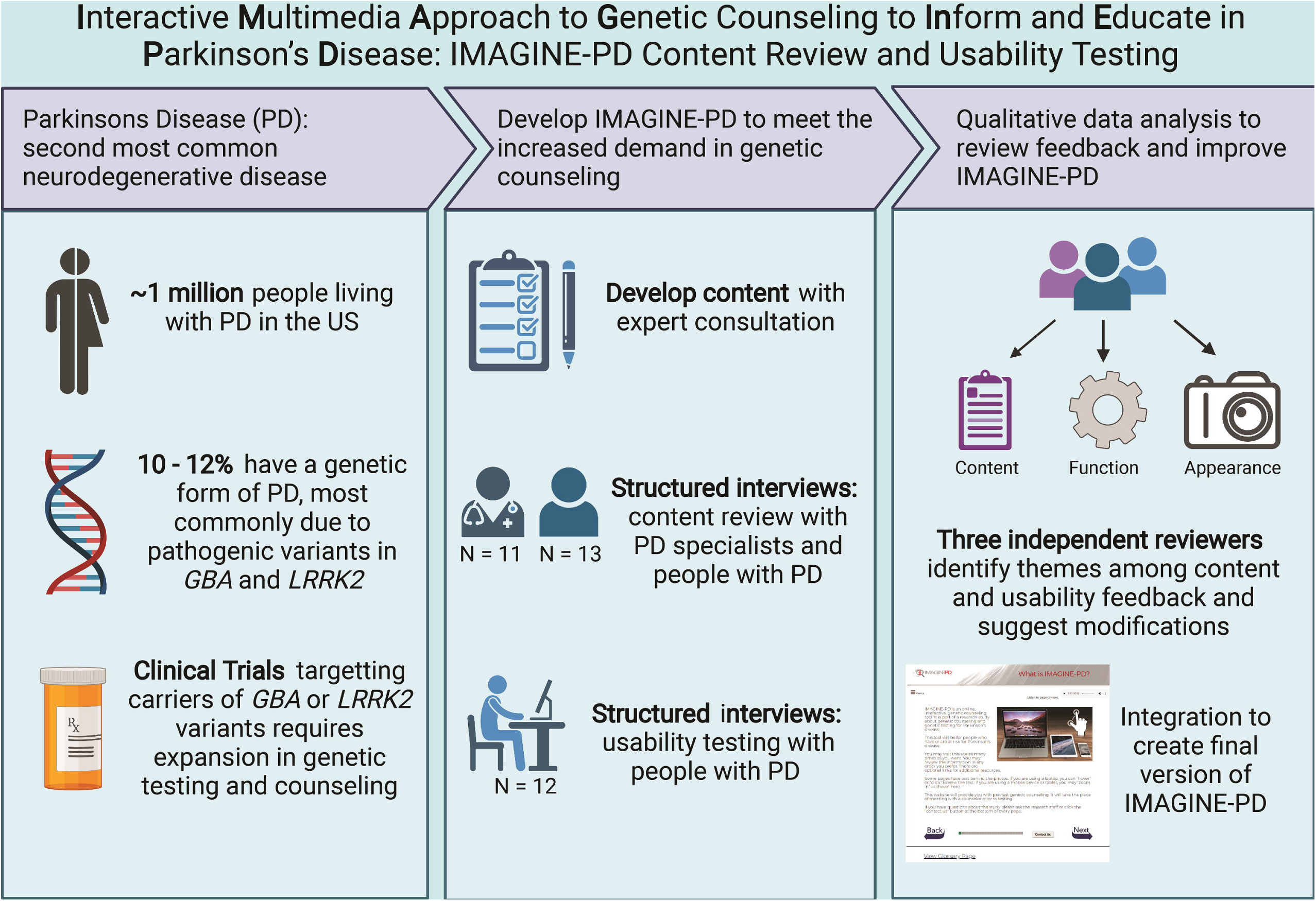
A. Parkinson’s’ disease (PD) patients and movement disorders specialists (MDS) self- reported use of the internet and email for health information (adapted from Baker et al. 2003).[21] B. PD patients and MDS self-reported knowledge in genetics, PD, and genetic testing (GT).

For each webpage participants were asked to rate the usefulness (1 being the least useful and 10 being the most useful). The average usefulness across all pages for PD patients was 8.49 (SD = 0.65) and for MDS was 8.84 (SD = 0.64). The results for usefulness for each page are found in **Supplemental Table 2** in the Supplement. Three domains of feedback were identified on user testing analysis: content, function, and appearance. Changes were made on 20 pages in response to content feedback, 4 pages in response to function feedback, and 4 pages in response to appearance feedback. A summary of the user testing qualitative analysis and recommendations for change are shown in **Table 2**.

**Table 2:** Summary of Phase 1 Content Review.

| Page | Content Feedback | Changes made |
| --- | --- | --- |
| 1. Introduction | Want more info on this page | Short summary was added on intro |
| 3. What is IMAGINE-PD | Not sure how the website is relevant to PD | Clarified study relevance to PD |
| 4. Summary | Wording is too technical | Simplified vocabulary |
| 5. What is PD | Wording is too technical | Simplified vocabulary |
| 6. How is PD diagnosed | Want more information on nonmotor symptoms | Information on nonmotor symptoms included |
| 7. What causes PD | Too much information on page | Information was cut down and simplified |
| 8. How is PD treated | Want less information on drugs, more information on other treatments | Drug section shortened and added other PD treatments |
| 9. What is genetics | Too much information on page | Information was shortened and simplified |
| 10. Basics of genetics | Too much information on page and too technical | Information was shortened and simplified |
| 11. What is GT | Wording is too technical and not sure how information is related to PD | Simplified vocabulary, clarified relevance |
| 12.5. Reasons for GT | Page should be included as separate page | Separate page was made for information |
| 13. Benefits and risks | Make it more related to PD and more information on benefits | Clarified PD relevance |
| 14. GT process | Too much detail | Simplified and shortened page |
| 15. How genetics affects PD | Too much information | Shortened page |
| 16. *GBA* and PD | Information too technical and want to know why this gene is being studied | Simplified vocabulary and clarified relevance of *GBA* |
| 17. *LRRK2* and PD | Information too technical and want to know why this gene is being studied | Simplified vocabulary and clarified relevance of *LRRK2* |
| 18. Other rare genes for PD | Language seems too technical, why only *GBA* and *LRRK2* being tested | Simplified language, provide more resources for additional information on other genes |
| 19. How do I get results | Language seems too technical, want more information | Clarified next steps and reviewed previously introduced topics to clarify information |
| 20. VUS and unexpected results | Wording too technical | Simplified language |
| 21. Implications & limitations | Confused about previously introduced topics | Added information to clarify |
| Page | Function Feedback | Changes made |
| 2. Instructions | Did not know that there was more information on the bottom and needed to scroll | Shortened page to minimize scrolling |
| 4. Summary | Picture was clickable but did not lead to any new page | Fixed error so picture was no longer clickable |
| 5. What is PD | Already knew information, wanted a skip function | Shortened page, so not as much time would be spent on it |
| 6. How is PD diagnosed | In page slides were confusing | Made in page slides more obvious |
| **Page** | **Appearance Feedback** | **Changes made** |
| 1. Introduction | Make title clearer and more obvious | Title was enlarged and bolded |
| 3. What is IMAGINE-PD | Use bullet points to make page easier to read | Used bullet points |
| 4. Summary | Picture was not relevant | Used a different picture |
| 12. When to consider GT | Did not like picture | Used a different picture |

At the conclusion, participants were asked to provide summary feedback. A summary of responses is found in **Supplemental Table 3** in the Supplement. Comments regarding the order of the presented information, the interactive tools such as the buttons, links, and menus, and overall comments are provided in **Supplemental Table 4** in the Supplement.

### Usability Testing

Participant demographics are summarized in **Table 1**. All participants reported using the internet or email within the past 12 months. One (8.3%) participant reported dial-up network use, 9 (75%) of participants reported broadband network use, and 7 (58%) of participants reported using smartphones, and 8 (67%) reported accessing the internet via a Wi-fi network. All participants reported using the internet to communicate with a health care provider, and 9 (75%) reported using the internet to search for health or medical information.

The same three themes were again identified on usability testing qualitative analysis: content, function, and appearance. Changes were made on 10 pages in response to content feedback, 6 pages in response to function feedback, and 7 pages in response to appearance feedback. A summary of the usability qualitative analysis and recommendations for change are shown in **Table 3**.

**Table 3:**
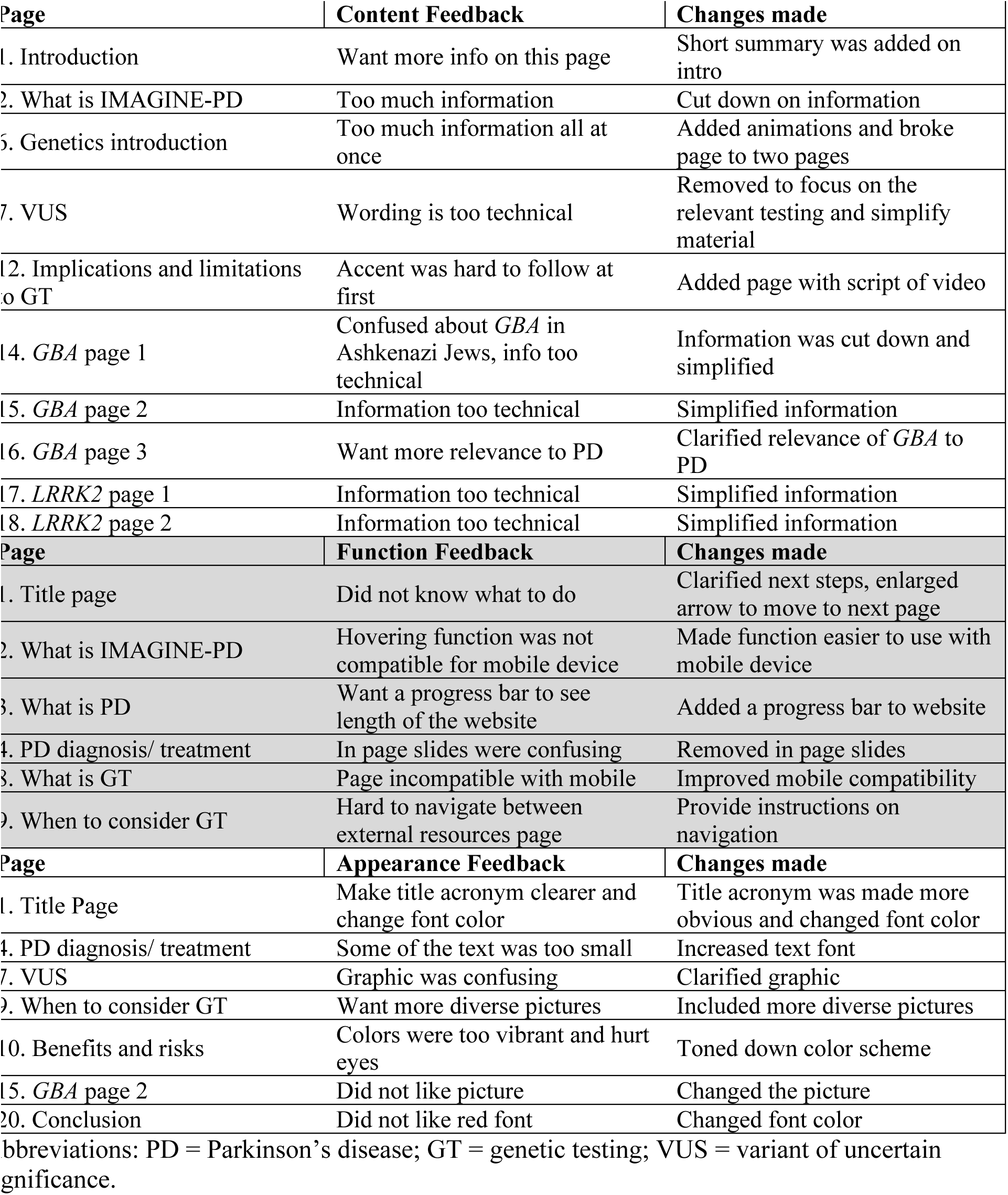
Summary of Phase 2 Usability Testing.

| Page | Content Feedback | Changes made |
| --- | --- | --- |
| 1. Introduction | Want more info on this page | Short summary was added on intro |
| 2. What is IMAGINE-PD | Too much information | Cut down on information |
| 6. Genetics introduction | Too much information all at once | Added animations and broke page to two pages |
| 7. VUS | Wording is too technical | Removed to focus on the relevant testing and simplify material |
| 12. Implications and limitations to GT | Accent was hard to follow at first | Added page with script of video |
| 14. *GBA* page 1 | Confused about *GBA* in Ashkenazi Jews, info too technical | Information was cut down and simplified |
| 15. *GBA* page 2 | Information too technical | Simplified information |
| 16. *GBA* page 3 | Want more relevance to PD | Clarified relevance of *GBA* to PD |
| 17. *LRRK2* page 1 | Information too technical | Simplified information |
| 18. *LRRK2* page 2 | Information too technical | Simplified information |
| Page | Function Feedback | Changes made |
| 1. Title page | Did not know what to do | Clarified next steps, enlarged arrow to move to next page |
| 2. What is IMAGINE-PD | Hovering function was not compatible for mobile device | Made function easier to use with mobile device |
| 3. What is PD | Want a progress bar to see length of the website | Added a progress bar to website |
| 4. PD diagnosis/ treatment | In page slides were confusing | Removed in page slides |
| 8. What is GT | Page incompatible with mobile | Improved mobile compatibility |
| 9. When to consider GT | Hard to navigate between external resources page | Provide instructions on navigation |
| Page | Appearance Feedback | Changes made |
| 1. Title Page | Make title acronym clearer and change font color | Title acronym was made more obvious and changed font color |
| 4. PD diagnosis/ treatment | Some of the text was too small | Increased text font |
| 7. VUS | Graphic was confusing | Clarified graphic |
| 9. When to consider GT | Want more diverse pictures | Included more diverse pictures |
| 10. Benefits and risks | Colors were too vibrant and hurt eyes | Toned down color scheme |
| 15. *GBA* page 2 | Did not like picture | Changed the picture |
| 20. Conclusion | Did not like red font | Changed font color |

Abbreviations: PD = Parkinson's disease; GT = genetic testing; VUS = variant of uncertain significance.

## Discussion

In this study, we evaluated a web-based genetics education tool for PD, using evidence-based research methods to refine the content and conduct usability testing. First, we developed content based on expert opinion and created a high-fidelity prototype. Next, we conducted user testing through structured interviews with MDS and PD patients to evaluate website content. Subsequently, we conducted usability testing via structured interviews with PD patients. Using qualitative data analysis in both phases, we identified three domains of feedback (content, function, and appearance), and addressed feedback by incorporating changes to IMAGINE-PD to create a final version.

The qualitative analysis in both phases identified important themes among the feedback and led to significant improvements in IMAGINE-PD. In user testing, content feedback pertained to the volume and level of detail of information (“too much information”), the complexity of the content (“wording is too technical”), and the focus of the content (“make it more related to PD”). To address these points, we made changes to focus and simplify the content and to use plain language at a Flesch-Kincaid reading level 9 or below. In usability testing, content feedback focused on the volume and level of detail of information and further refinements were made. Key points of feedback about website function included requests for a progress bar and mobile compatibility, which were both addressed for the final version. Feedback about appearance included font color and size, as well as picture choice. Final pictures were selected to ensure representation of individuals of diverse race, ethnicity, age, and sex.

Some studies in PD and Alzheimer’s disease (AD) have employed the use of alternative media forms as pre-test education tools. For instance, the PDGENEration study, which offers genetic testing and counseling for PD, provides a pre-test education tool that is a pre-recorded video covering essential topics in PD genetics and genetic testing that was created by experts in neurogenetics and genetic counseling for PD (NCT04057794). Additionally, the Alzheimer’s Prevention Initiative, which conducts *APOE* testing in cognitively unimpaired people over 60 years old, utilizes a self-directed learning technique providing a brochure and a video covering content typically addressed in a pre-test counseling session coupled with multiple choice questions to reinforce learning.[24] To our knowledge, neither approach has undergone usability testing to incorporate the input of the end-user as we demonstrate in this study.

Beyond neurogenetics, these results can also be viewed in the context of alternative genetic education tools for inherited cancer syndromes, where more robust efforts to develop alternate education and disclosure methods are underway. In the Communication and Education in Tumor Profiling (COMET) and the Returning Genetic Research Panel Results for Breast Cancer Susceptibility (RESPECT) studies online educational tool for pre-test education was developed using the same evaluation methods used in this study, serving as a guide for user testing and usability testing of IMAGINE-PD.[25, 26] Furthermore, in the REACH3 and the Study of an eHealth Delivery Alternative for Cancer Genetic Testing for Hereditary Predisposition in Metastatic Cancer Patients (eREACH) studies, web-based alternatives to traditional provider-mediated counseling and results disclosure are being evaluated. The outcomes of these studies will be informative for understanding the use of web-based genetic education tools even beyond inherited cancer syndromes. However, differences in the target populations, genetic testing performed, and the implications of the genetic test results between IMAGINE-PD and the COMET, RESPECT, and REACH studies necessitated the rigorous user testing and usability testing that we report here.

This study has several strengths that should be noted. First, the content for IMAGINE-PD was developed by experts in genetic counseling in PD, neurogenetics experts, and movement disorder physicians. Second, the user testing includes the typical referring providers for neurogenetic services for PD patients (movement disorder physicians), and the intended end-user (PD patients). Third, this evaluation followed the DHHS guidelines for user testing and usability testing, nearly doubling the recommended sample size in each phase for this type of research.

Some limitations of this study should be acknowledged. First, the content of IMAGINE- PD is focused on targeted variant testing in *GBA* and *LRRK2*, limiting its scope and generalizability to other PD genetic testing. This was intentional to allow IMAGINE-PD to serve as a follow-up for results disclosure for the UPenn MIND study.[20] The MIND study conducts *GBA* and *LRRK2* targeted variant screening in all PD patients seen at the PDMDC at UPenn using the same genetic tests for all involved study participants. As a result, the pre-test education in the IMAGINE-PD tool deviates from a typical pre-test genetic counseling session that would involve obtaining a family history and making decisions about test choice (family variant testing, multi-gene panel testing, exome, or genome sequencing). Instead, this is a scalable approach to screen everyone in the UPenn PD clinic for variants within two most common genes associated with PD, to identify potentially eligible participants for clinical trials enrolling carriers of variants in *GBA* or *LRRK2*. During a separate disclosure visit for *GBA* or *LRRK2,* a personal and family history could be reviewed in detail, and additional testing could be pursued afterwards, if indicated. Additionally, specialized content could be developed and added to subsequent versions of IMAGINE-PD to accommodate other specific types of diagnostic genetic testing. Second, all participants (physicians and patients) had a high level of education and experience with technology and computer use, and interest in using an online pre-test education tool, which probably does not capture the breadth of PD patients and may overestimate the user experience. Ongoing evaluation of this tool will be necessary to determine which patients will be able to successfully use an online educational platform and who would be better served by other methods such as in-person or live telemedicine counseling with a genetic counselor.

Additionally, supplementary content could be developed to address deficits in patient or family-member comprehension, low-literacy or education level, identifying potential psychosocial concerns to prompt additional counseling, or comfort with technology. Making this tool accessible while in clinic on a smart-phone, tablet, or computer screen may help to address limitations in access to technology.

In summary, we present our findings from user testing and usability testing for the development of IMAGINE-PD, an online pre-test genetic education tool. We describe a phased review and iterative process of refining the content, appearance, and functionality based on expert review as well as physician and patient feedback according to DHHS guidelines. The final version, which is available at http://www.pennmedicine.org/imaginepd, will undergo further evaluation to compare it to standard tele-genetic counseling with a genetic counselor (NCT04527146) measuring satisfaction, impact, and knowledge. As a virtual learning tool accessible anywhere by internet, IMAGINE-PD has potential to improve access to neurogenetic services for PD patients interested in learning about their eligibility for *LRRK2* or *GBA* directed clinical trials.

## Data Availability

All data produced in the present study are available upon request to the authors

http://www.pennmedicine.org/imaginepd

## Acknowledgements

The authors would like to acknowledge our patients for their generous participation in this study, the clinical research associates at the University of Pennsylvania, and the expert reviewers of the preliminary material. We would also like to thank Dr Judy Shea, PhD at the University of Pennsylvania who provided guidance on qualitative data analysis, Susan Paolin who created the animations, and Stephen Durborow who created the website. Support and funding were provided by the Penn Center for Precision Medicine, the University of Pennsylvania Department of Neurology, The Parkinson Council, and the Parker Family Chair (ACP).

## Conflict of Interest

The authors have no conflict of interest to report

## Figure Legends

**Includes:**

Supplemental Table 1.

Supplemental Table 2.

Supplemental Table 3.

Supplemental Table 4.

Supplemental Questionnaire 1.

Supplemental Questionnaire 2.

**Supplemental Table 1.**
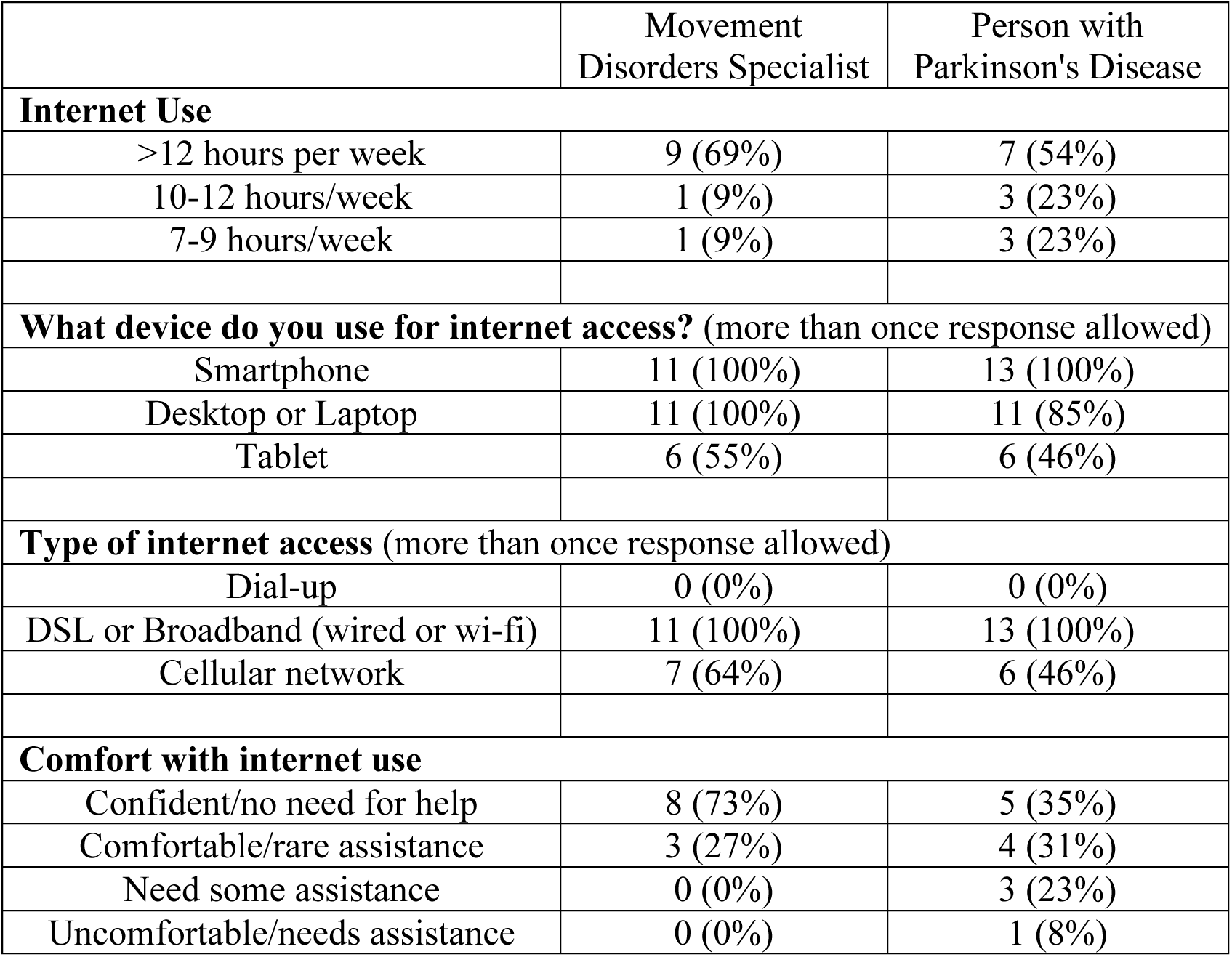
Internet use reported by movement disorders specialists (N=11) and people with Parkinson’s disease (N=13) in the content review (Phase 1). Available data presented as precent of total cohort.

**Supplemental Table 2.**
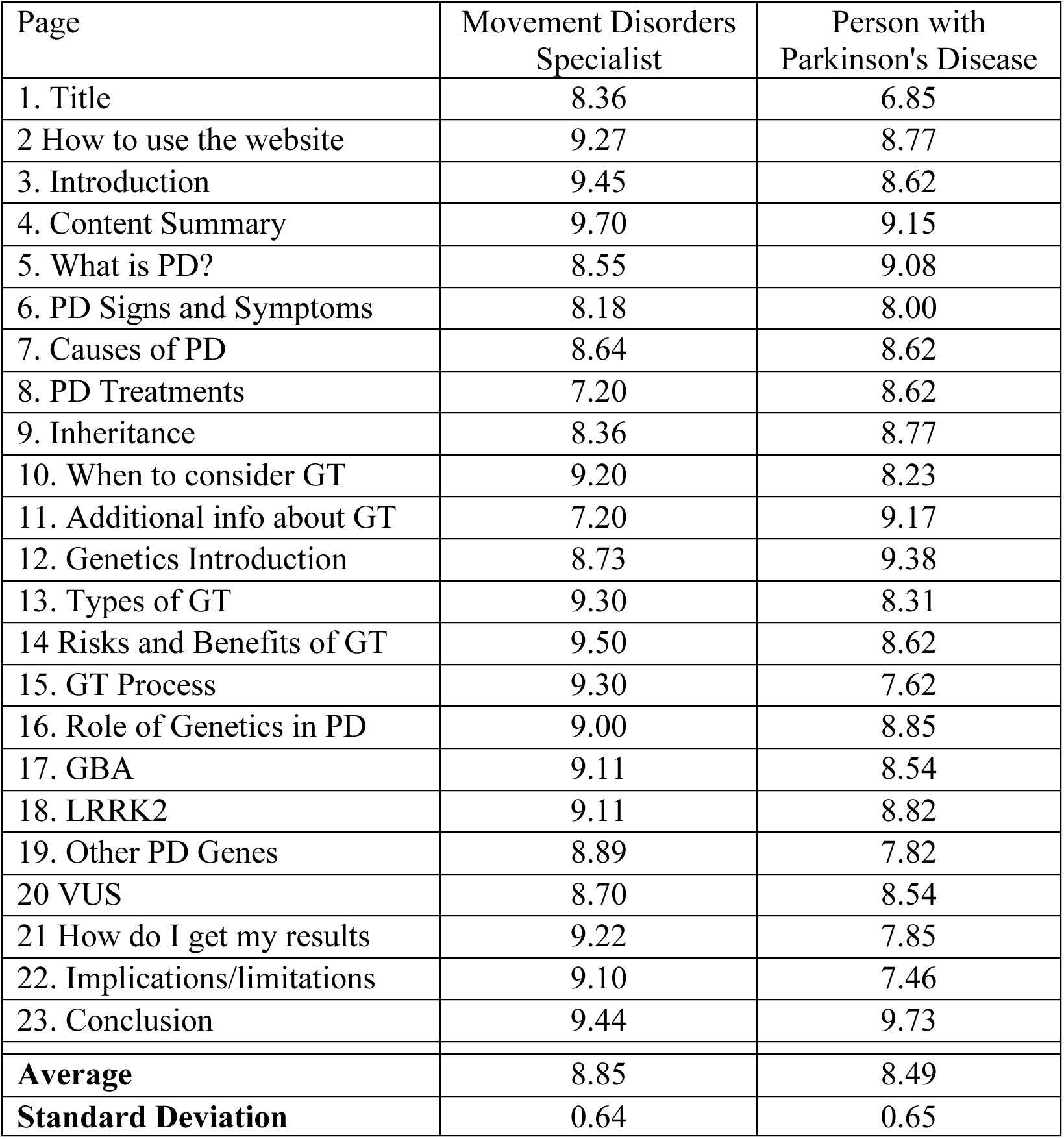
Average usefulness scale for each page reported by movement disorders specialists (N=11) and people with Parkinson’s disease (N=13) in the content review (Phase 1). Usefulness is reported from 1-10 with 1 being the lowest and 10 being the highest. PD = Parkinson’s disease. GT = Genetic Testing. VUS = Variant of uncertain significance.

**Supplemental Table 3.**
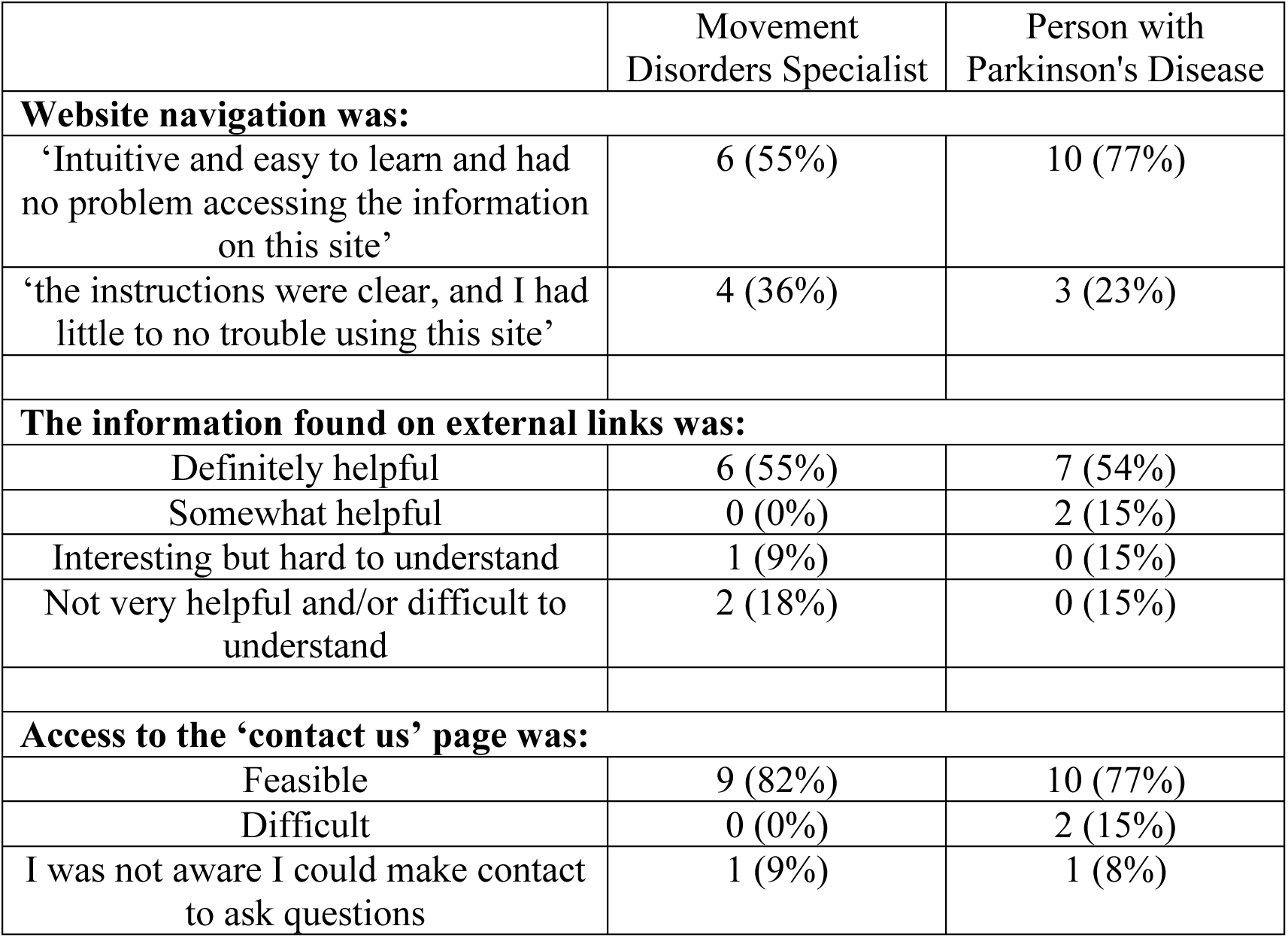
Summary feedback reported by movement disorders specialists (N=11) and people with Parkinson’s disease (N=13) in the content review (Phase 1). Available data presented as percent of total cohort.

**Supplemental Table 4.**
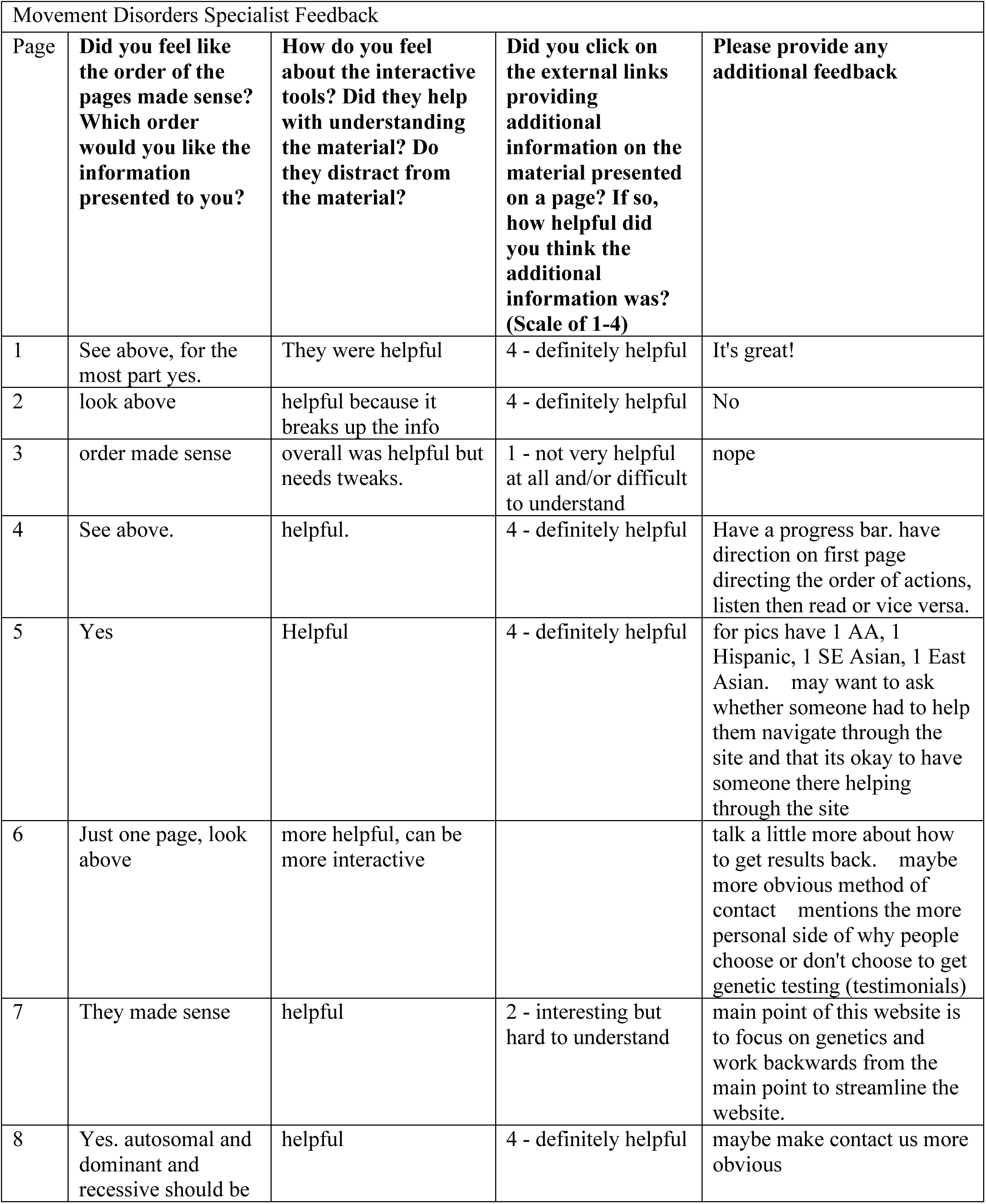
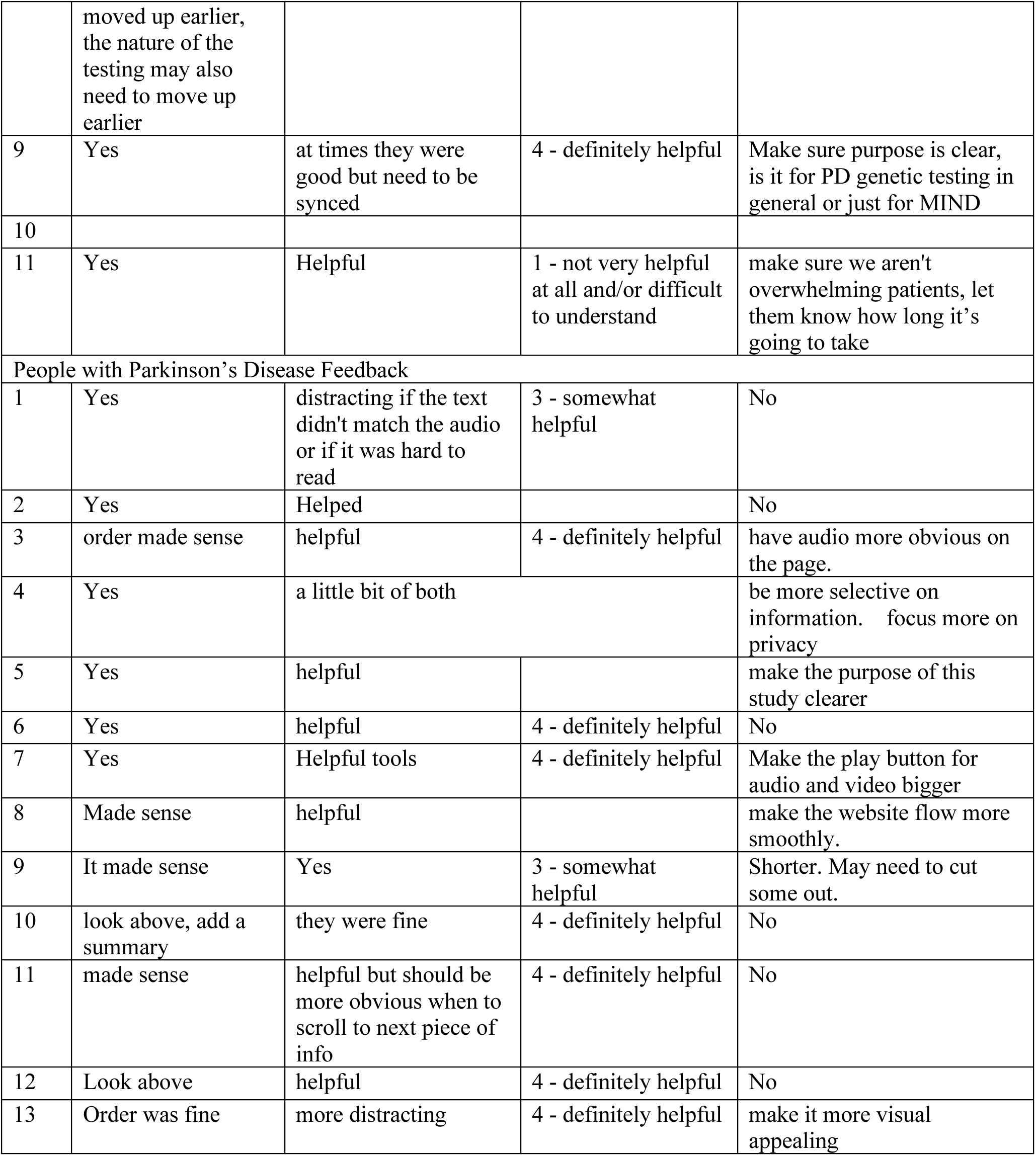
Final remarks from movement disorders specialists (N=11) and people with Parkinson’s disease (N=13) in the content review (Phase 1).

## Content Review Questions

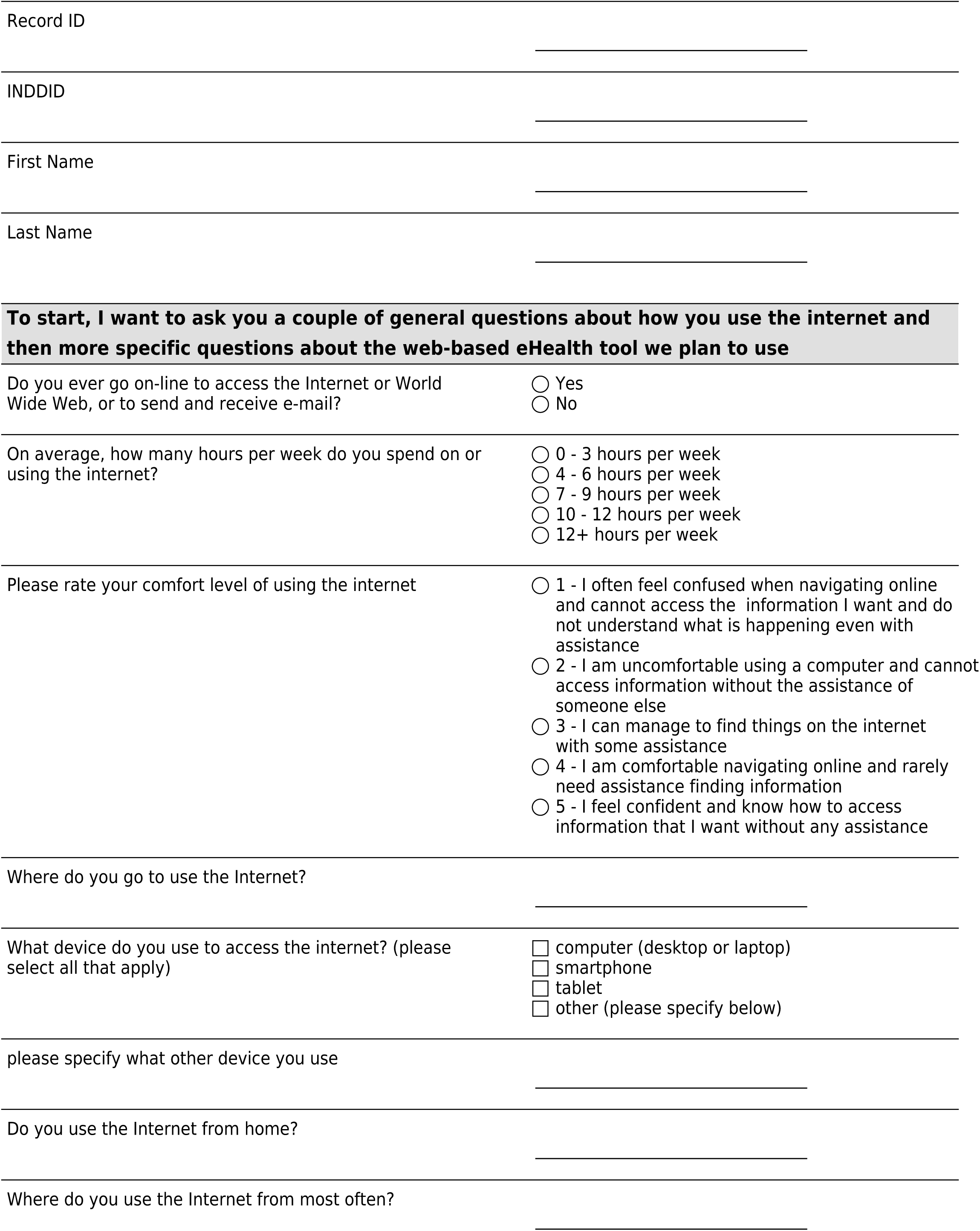
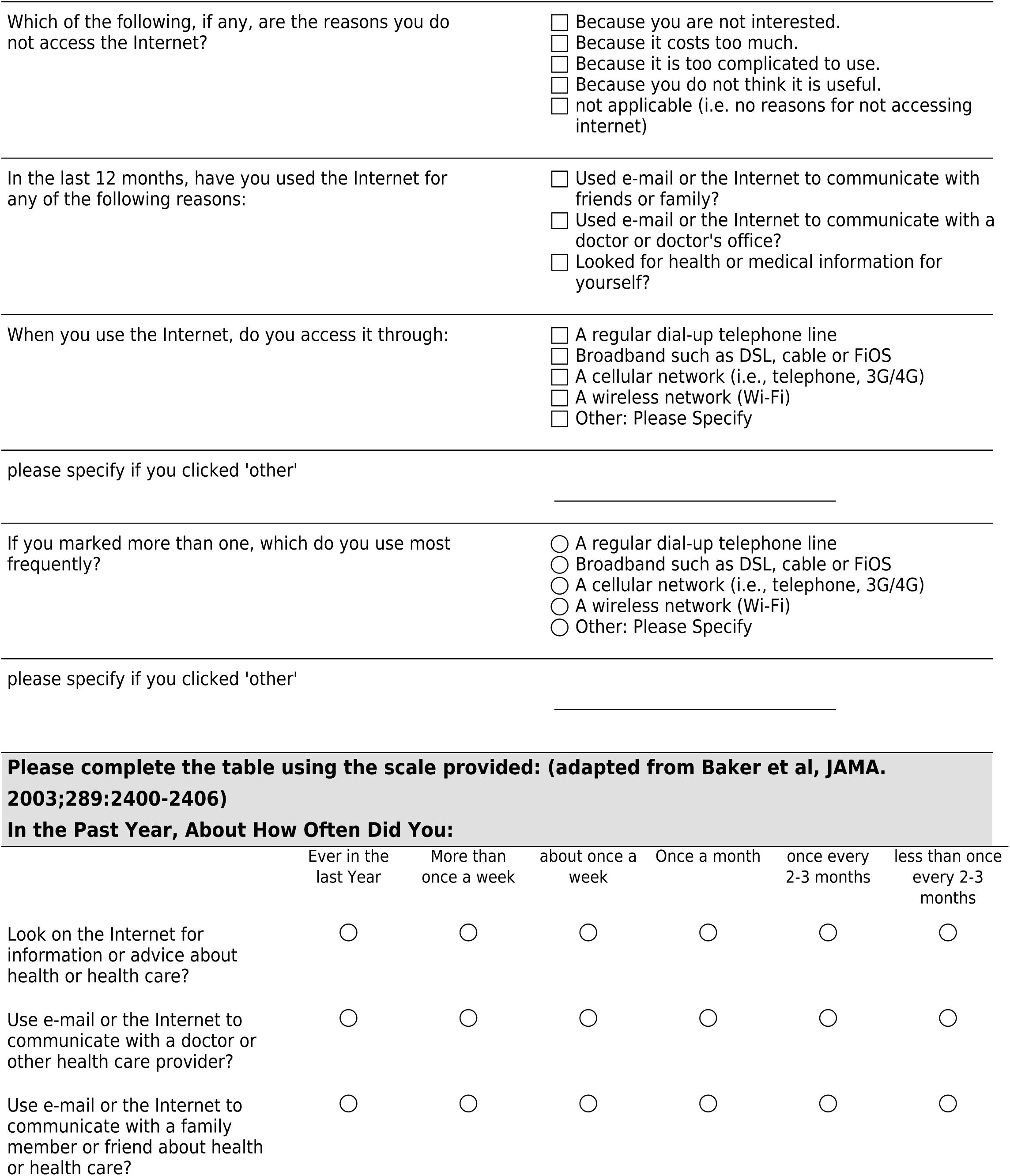
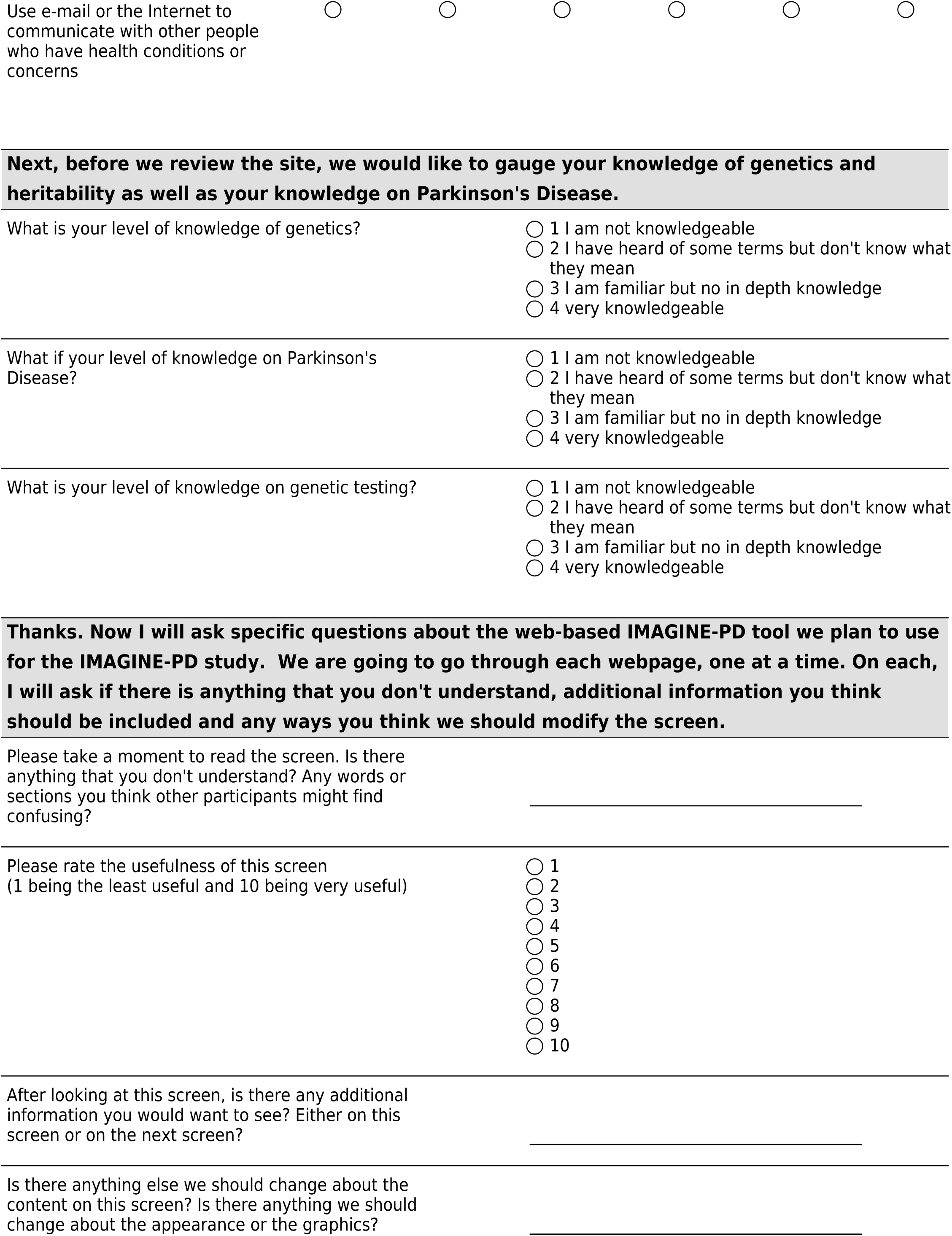
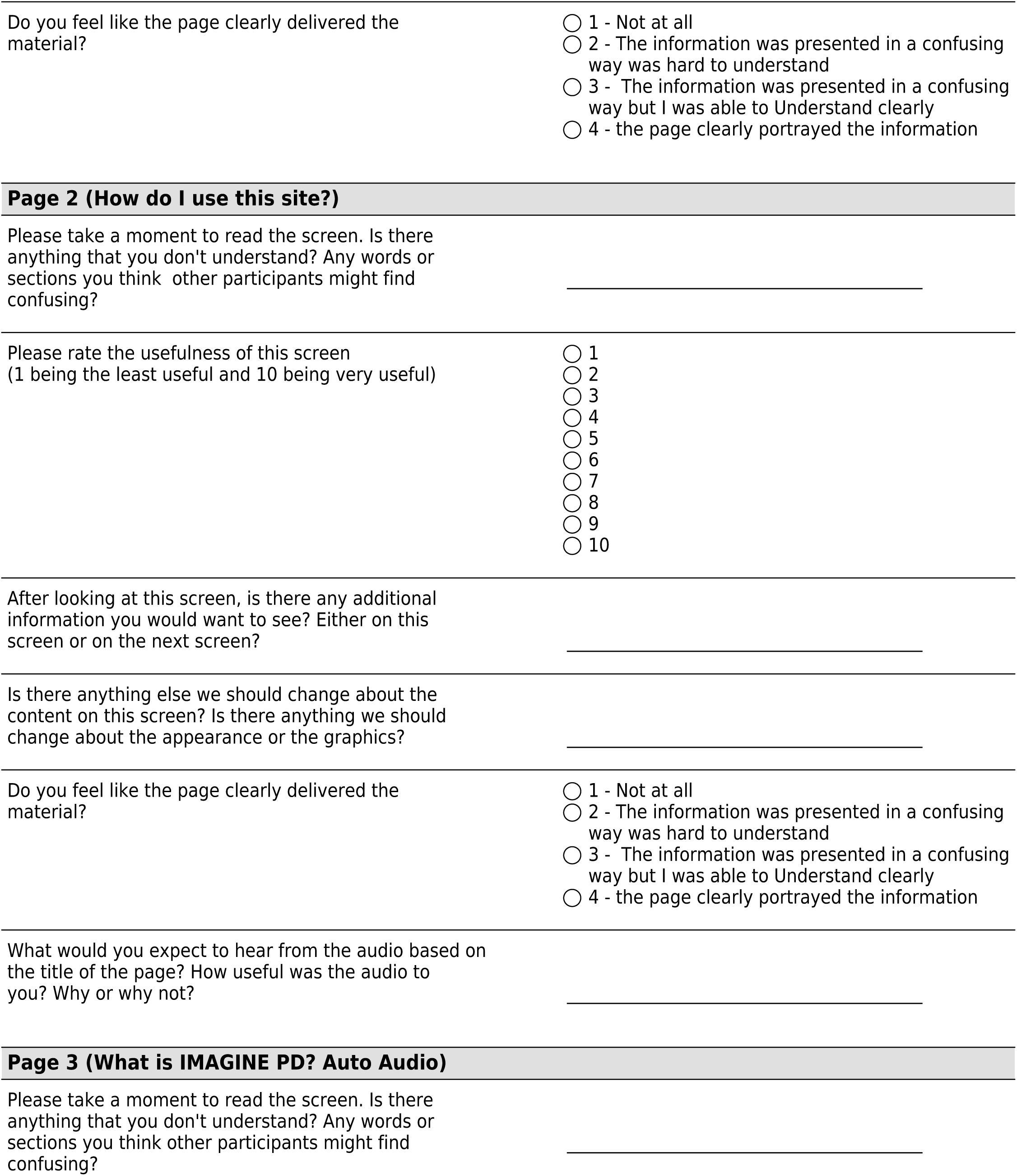
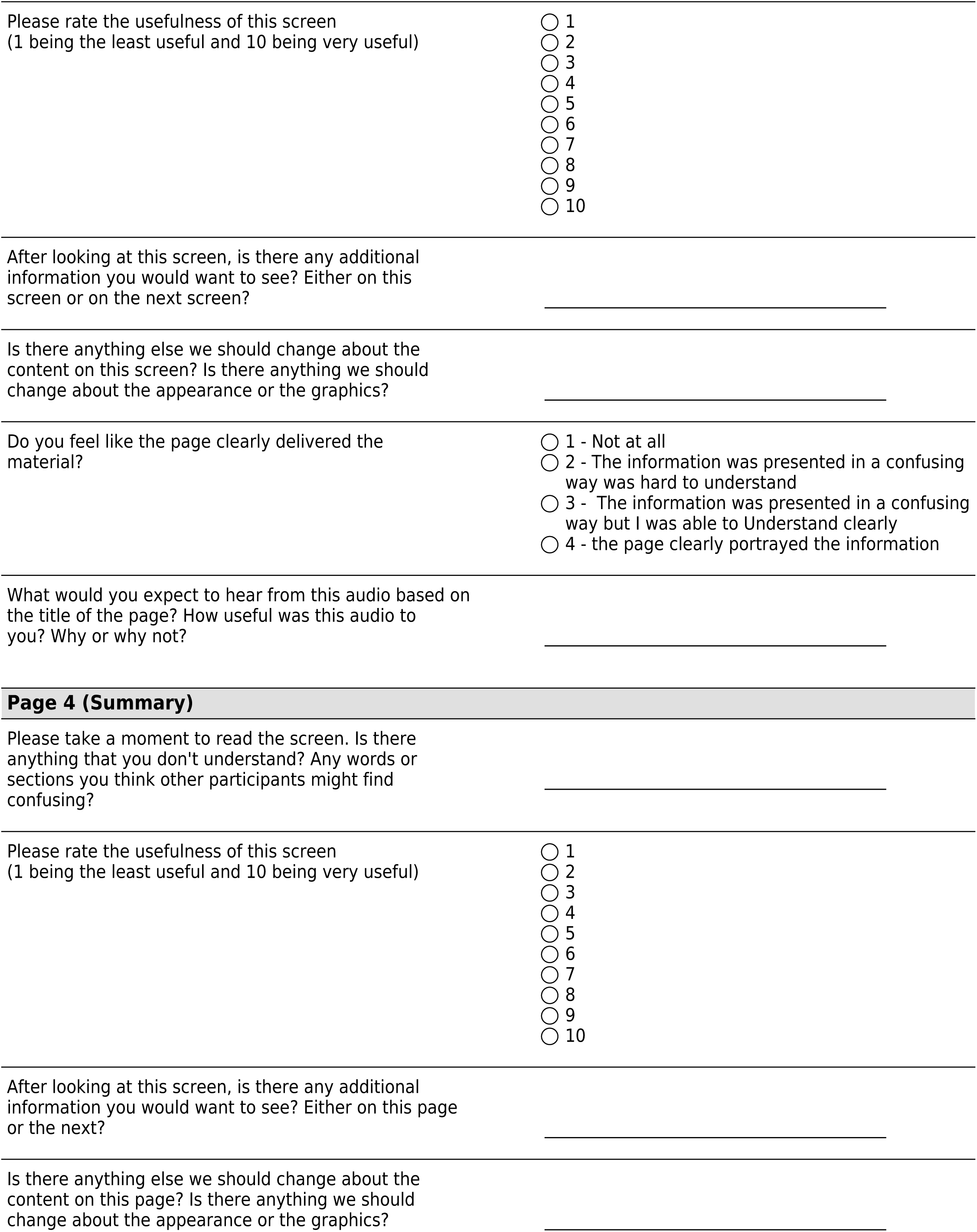
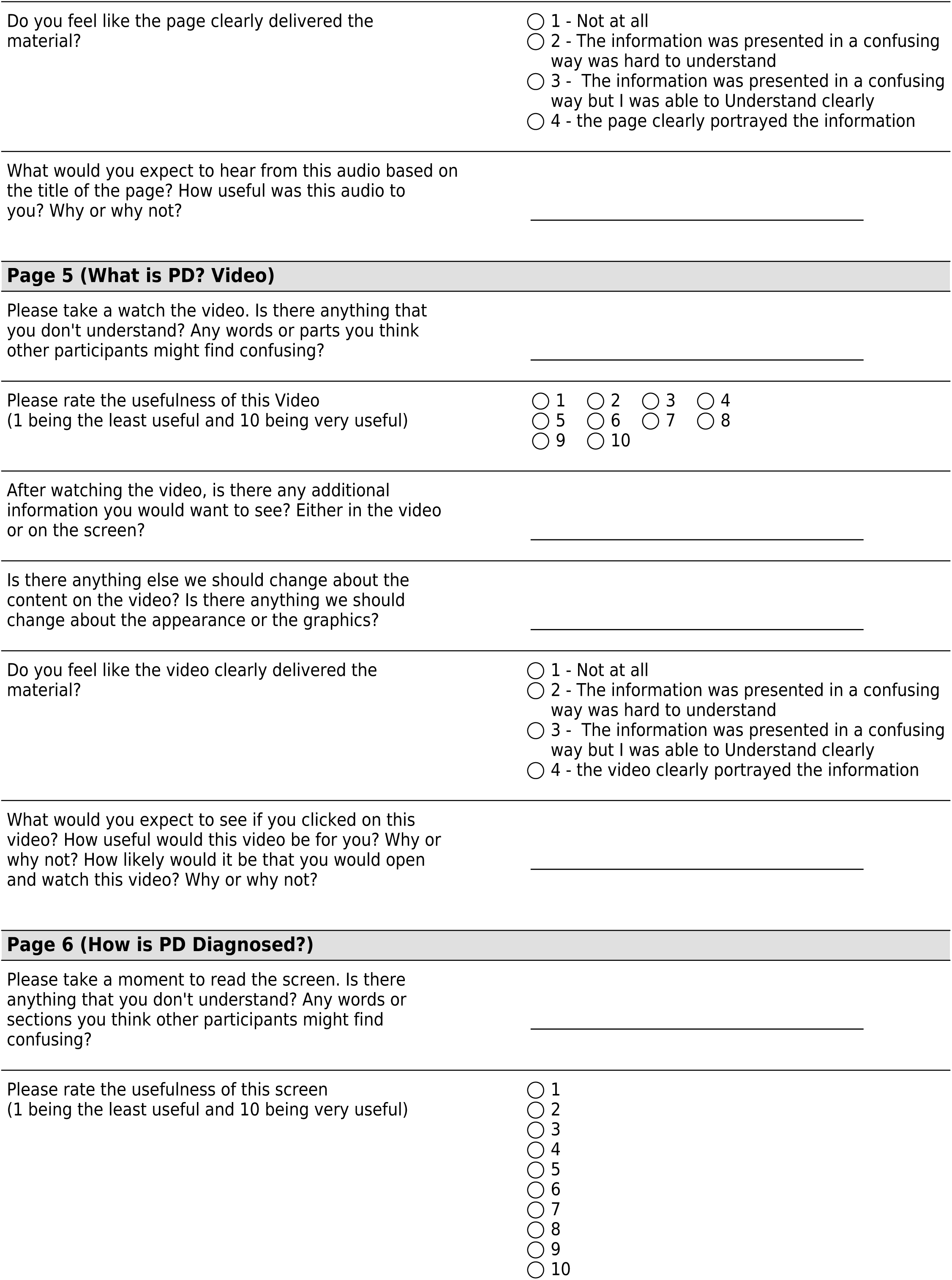
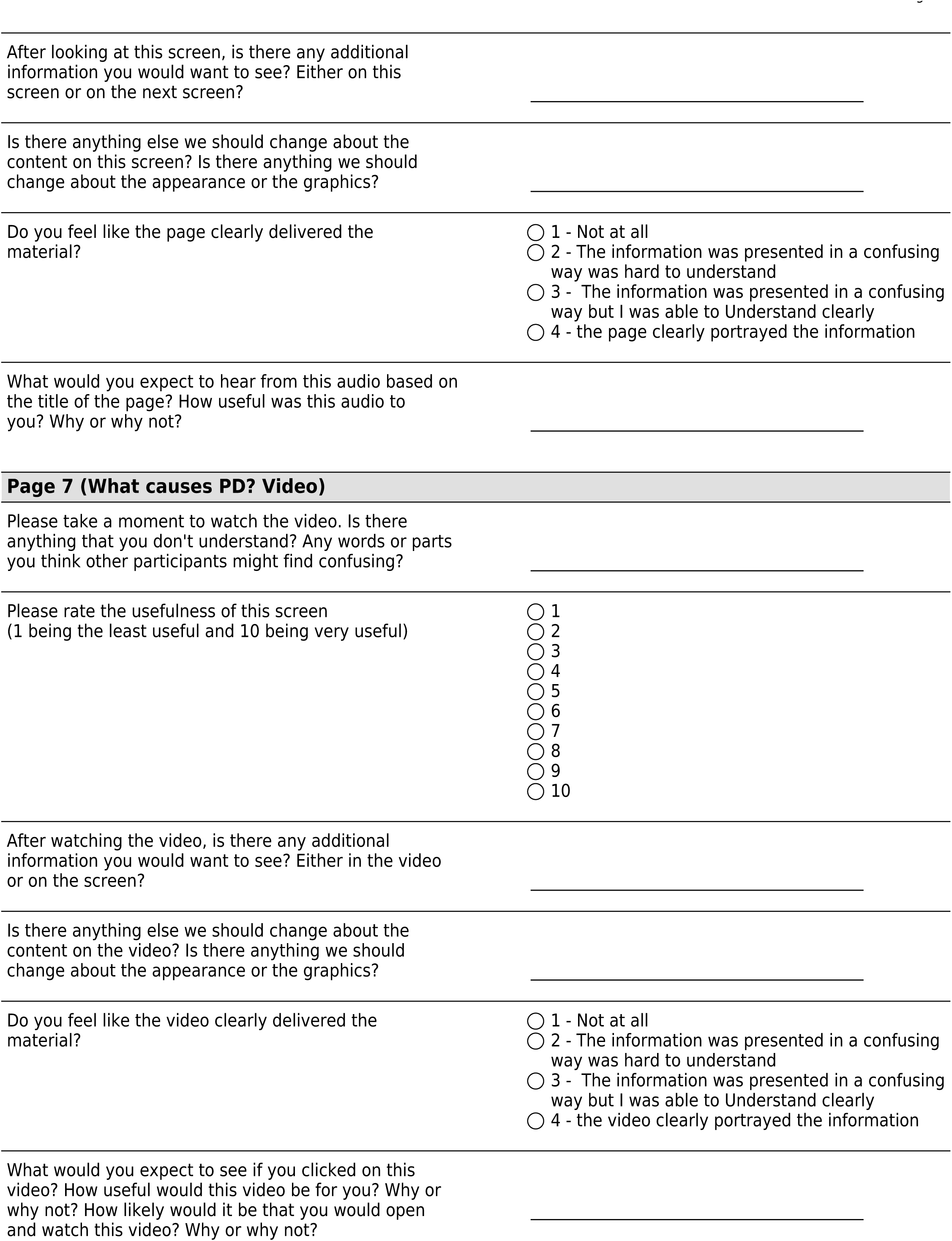
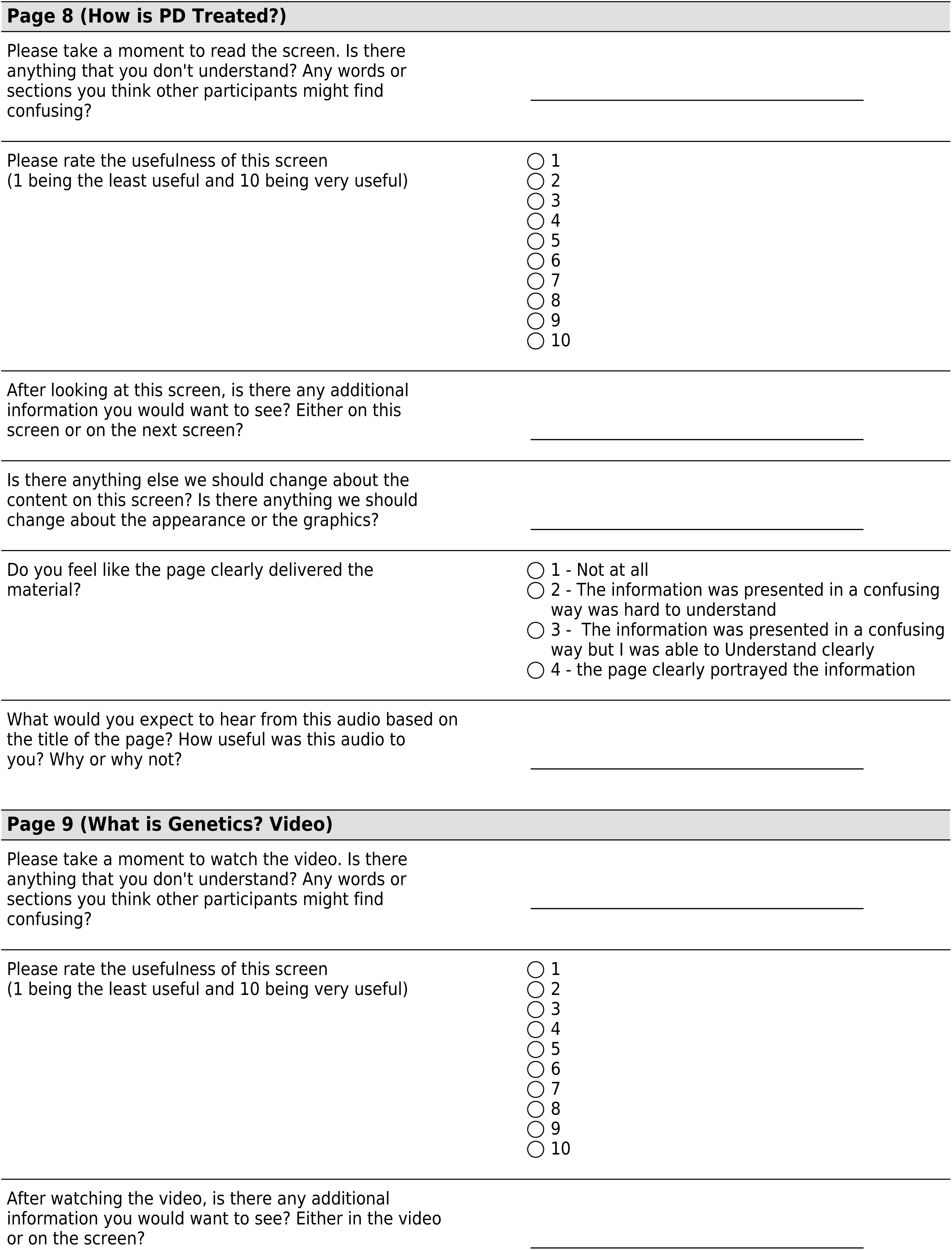
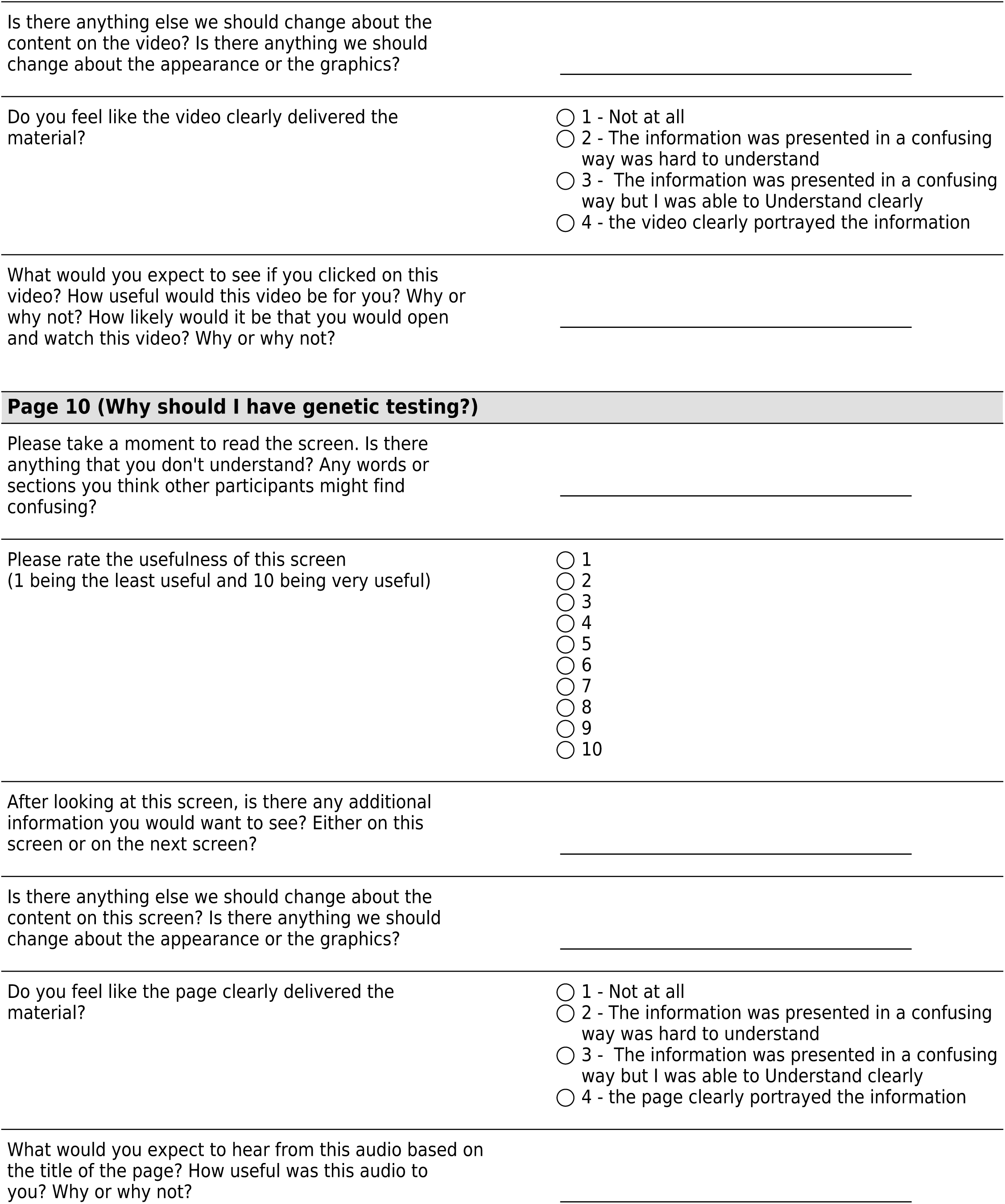
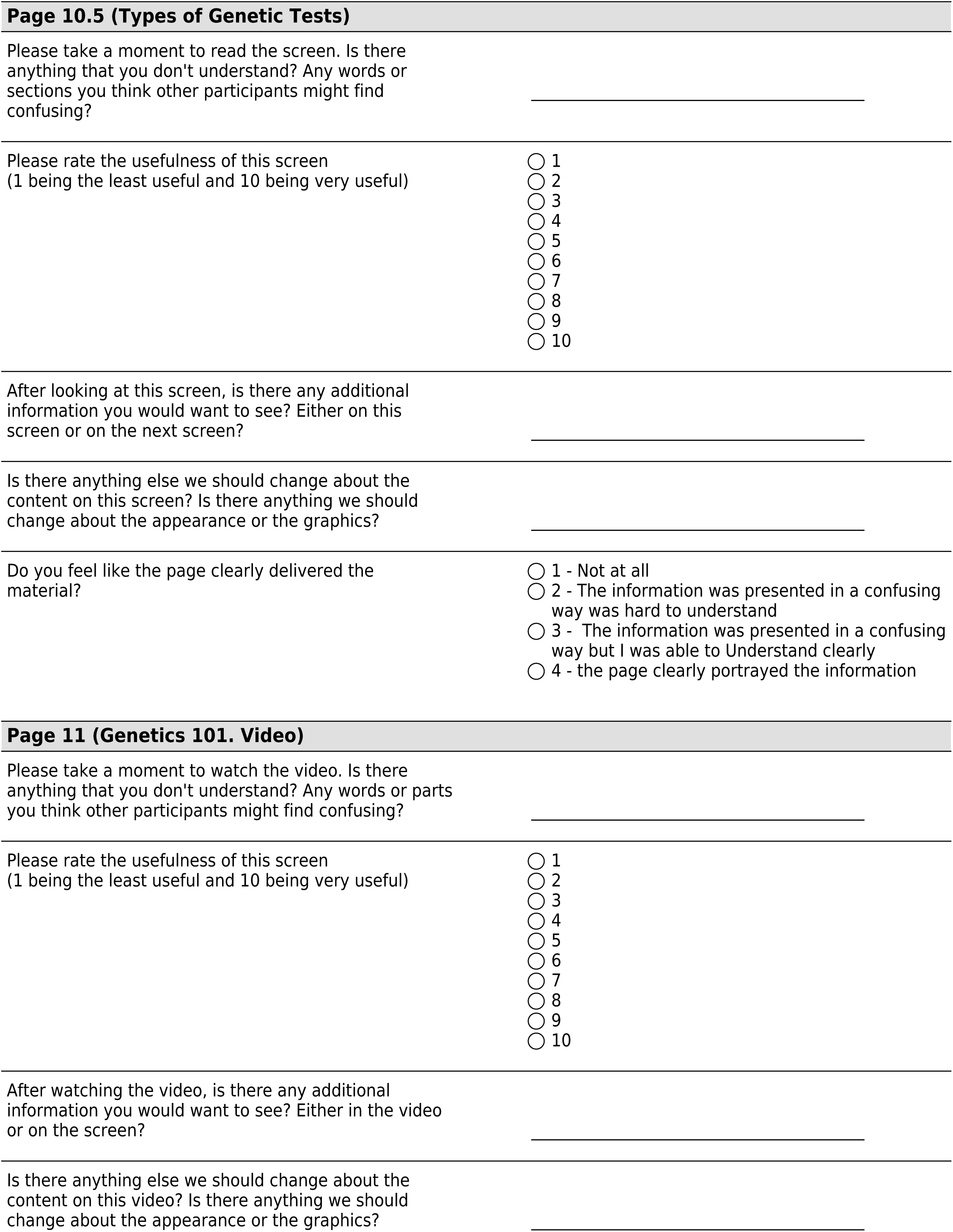
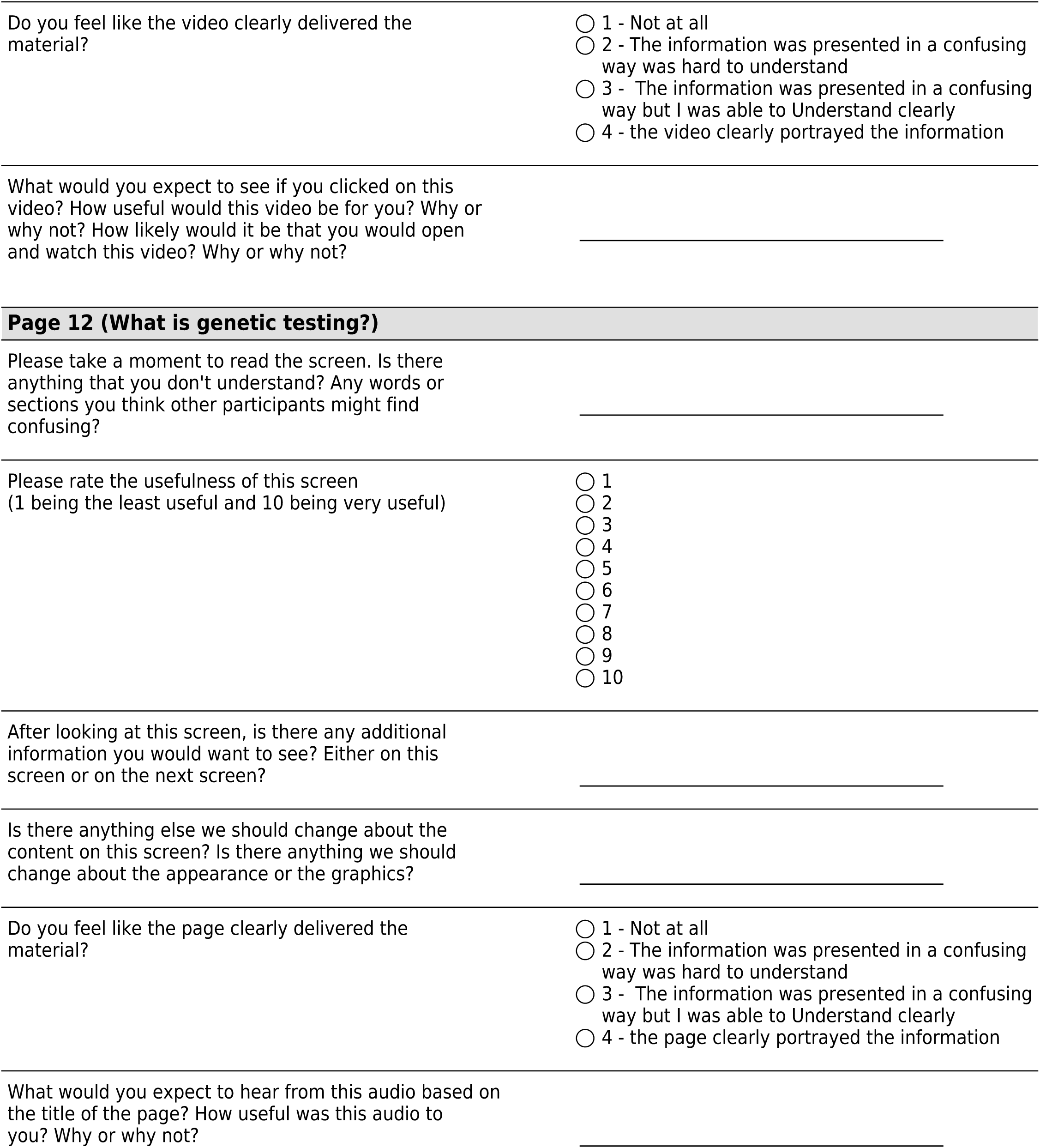
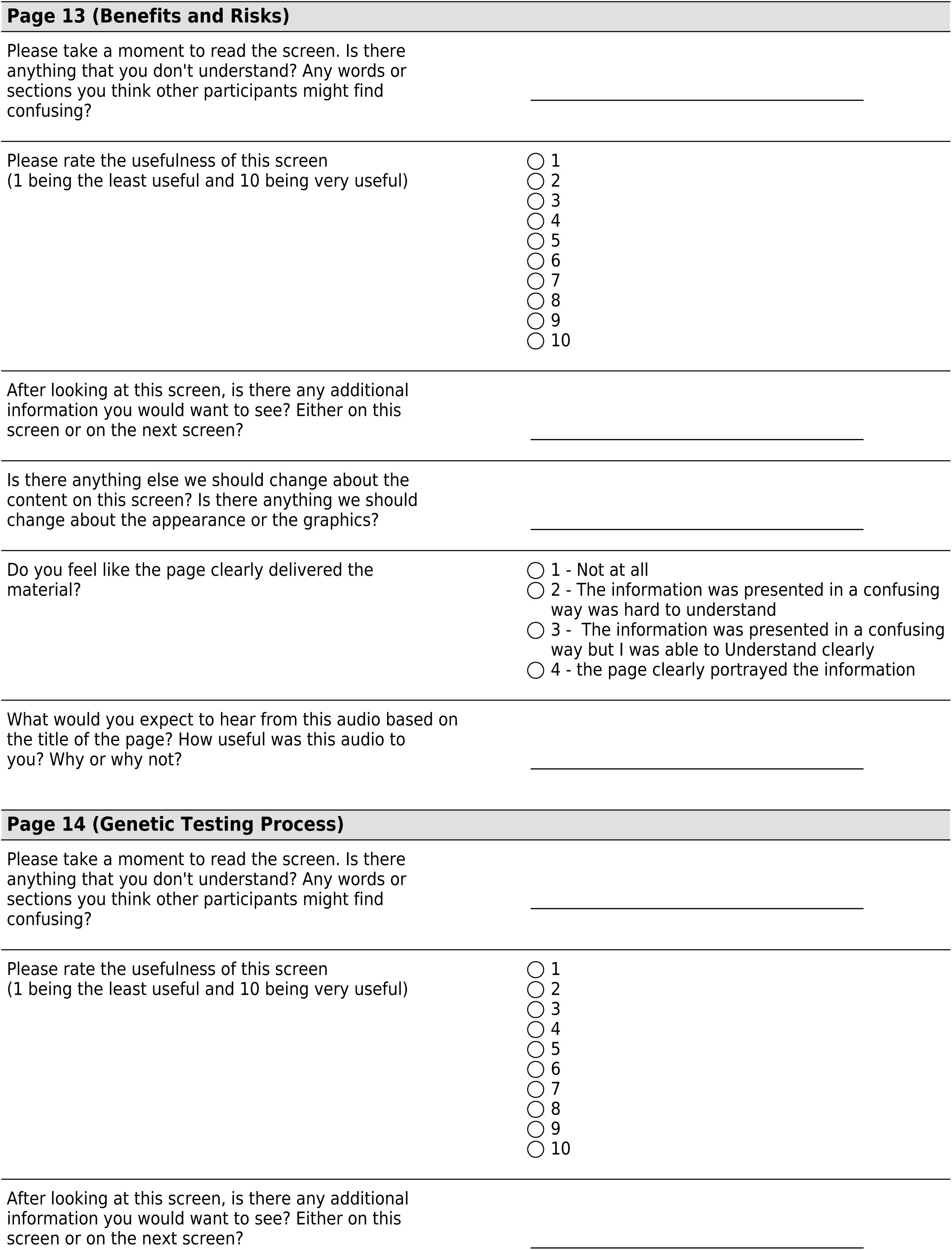
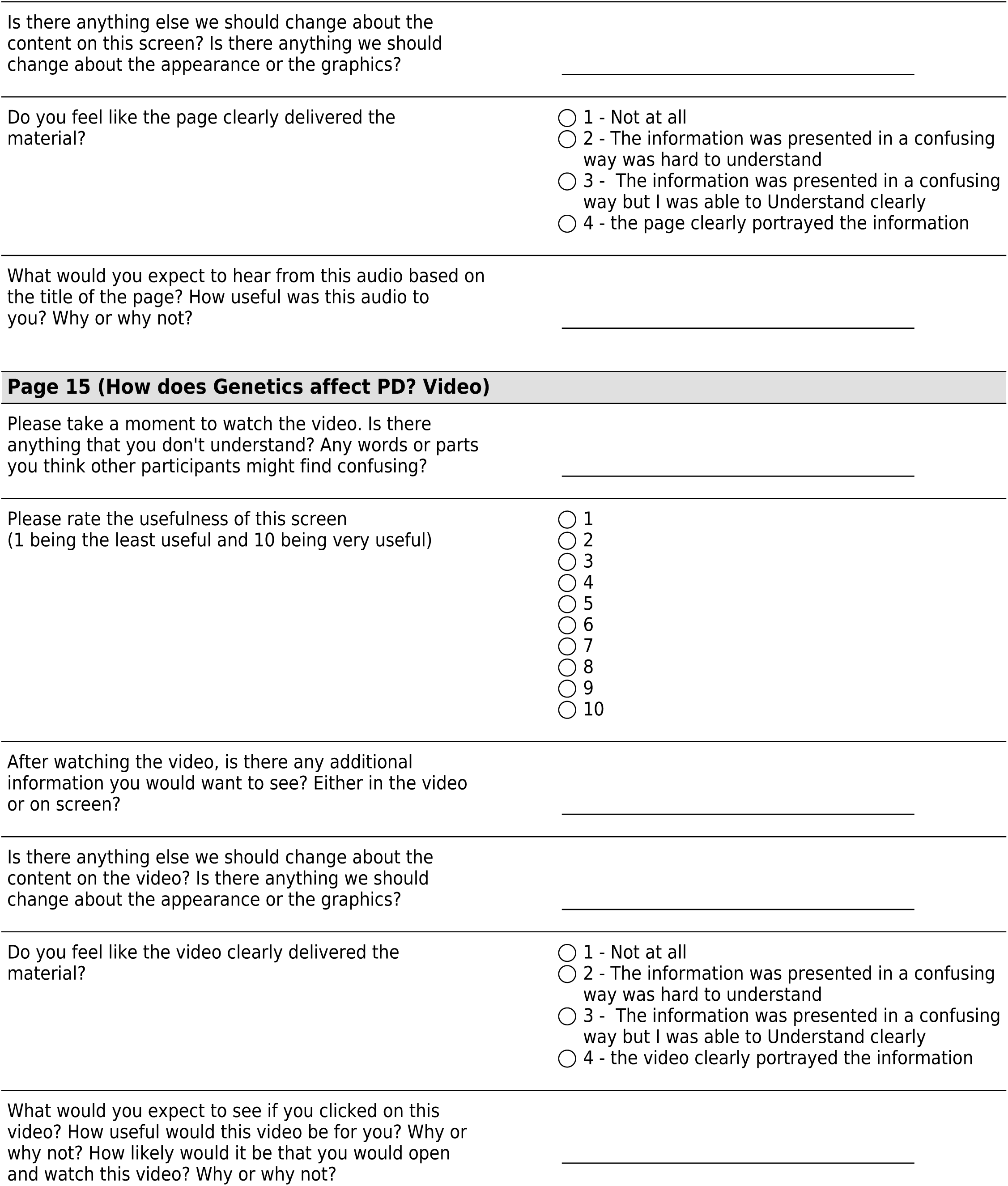
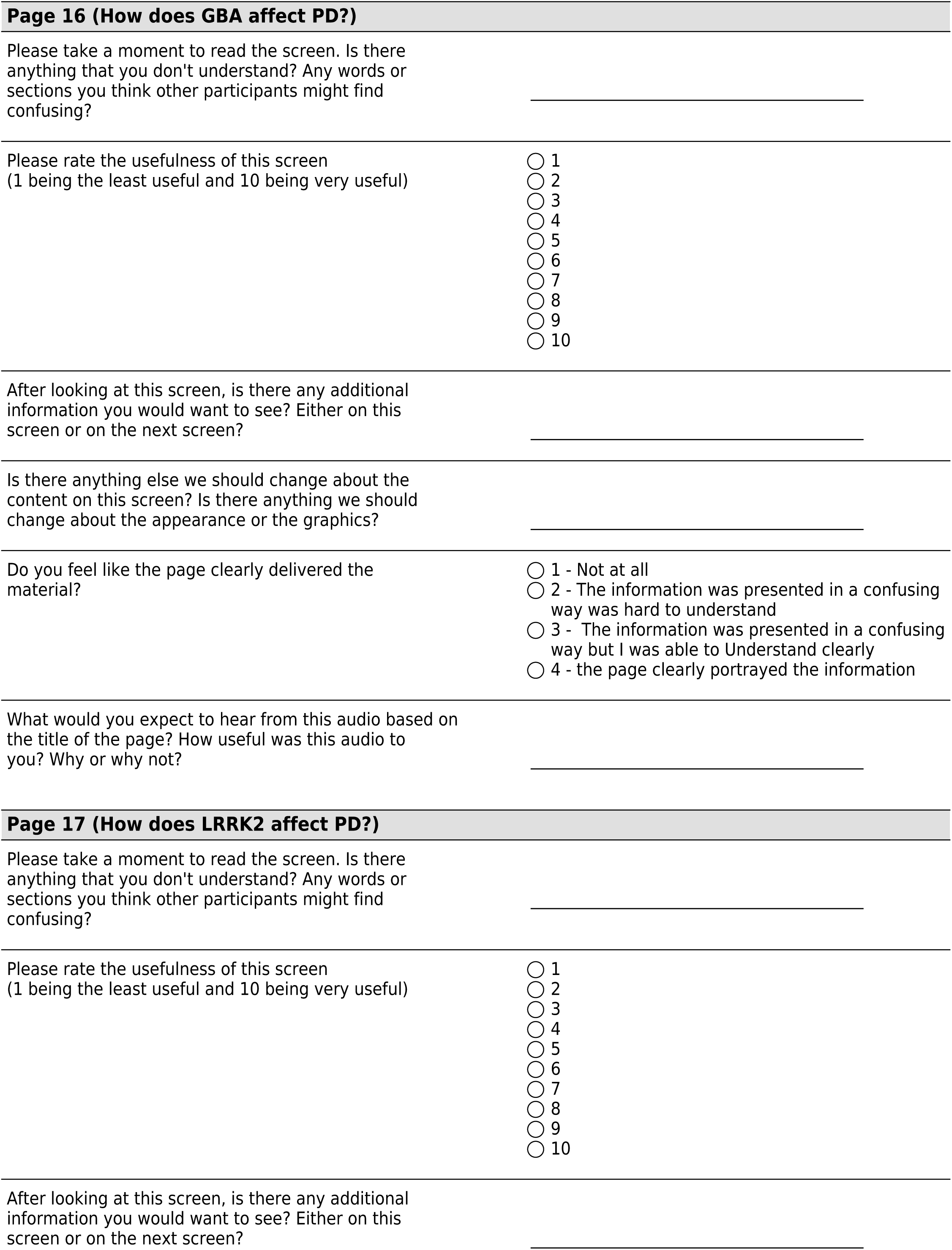
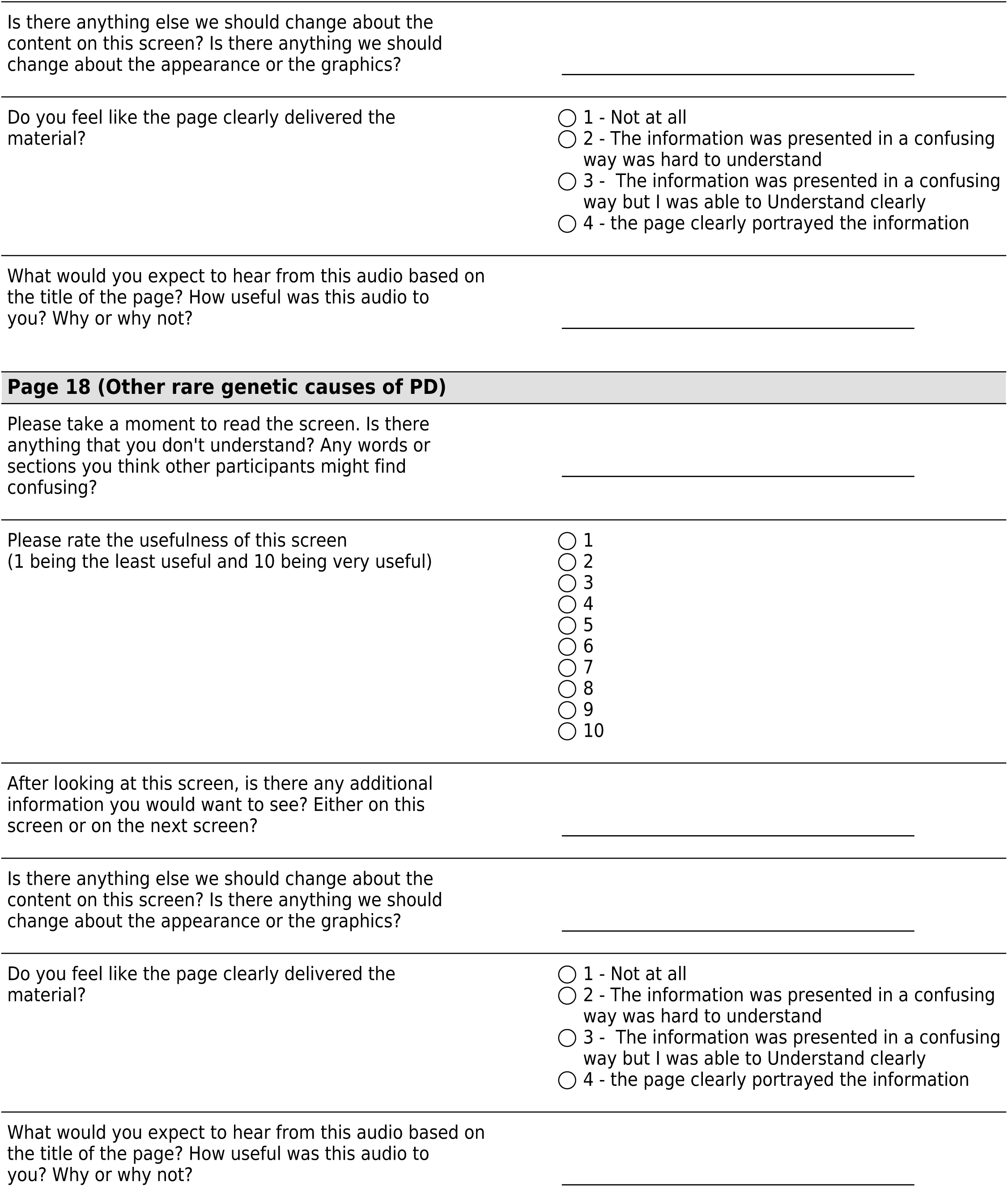
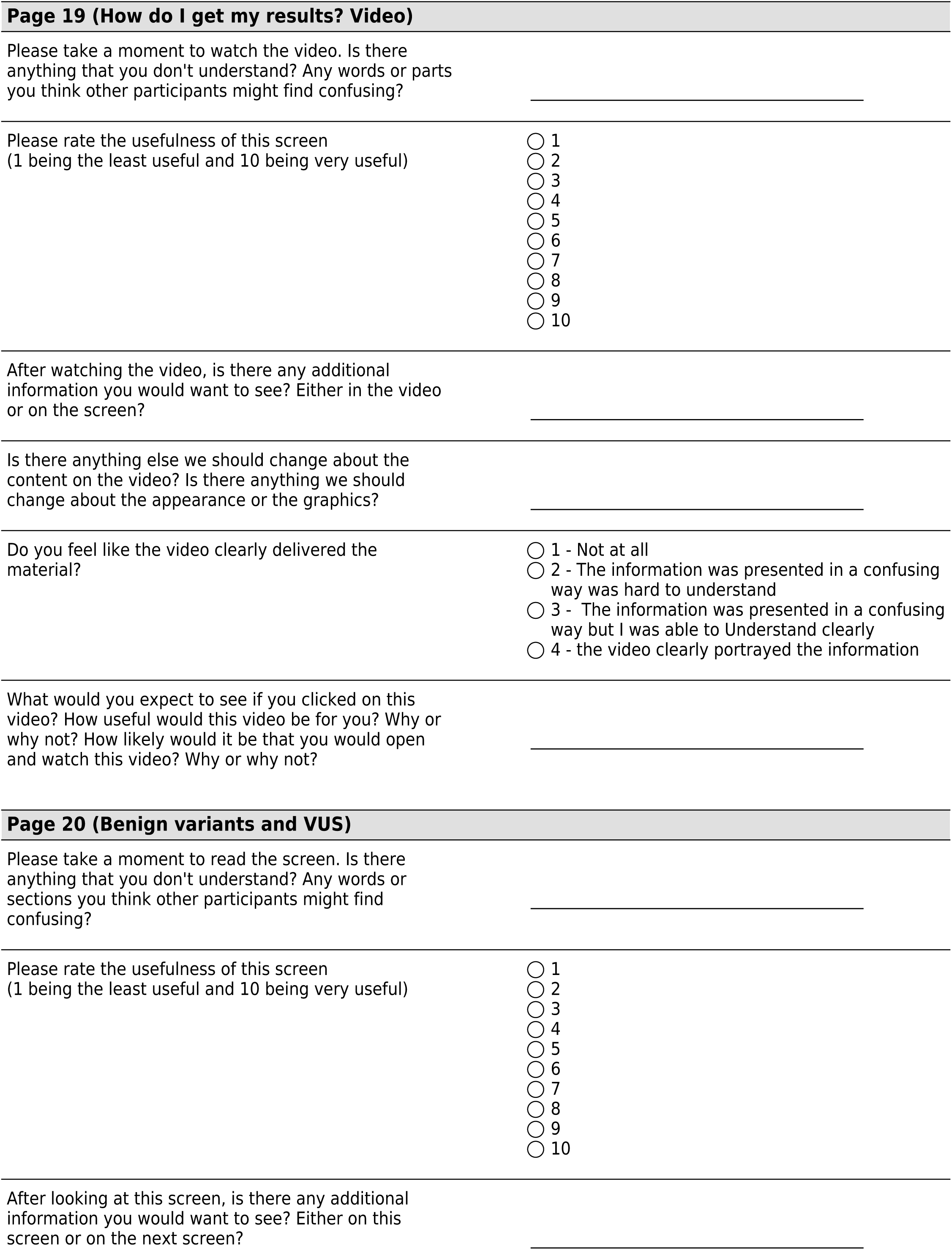
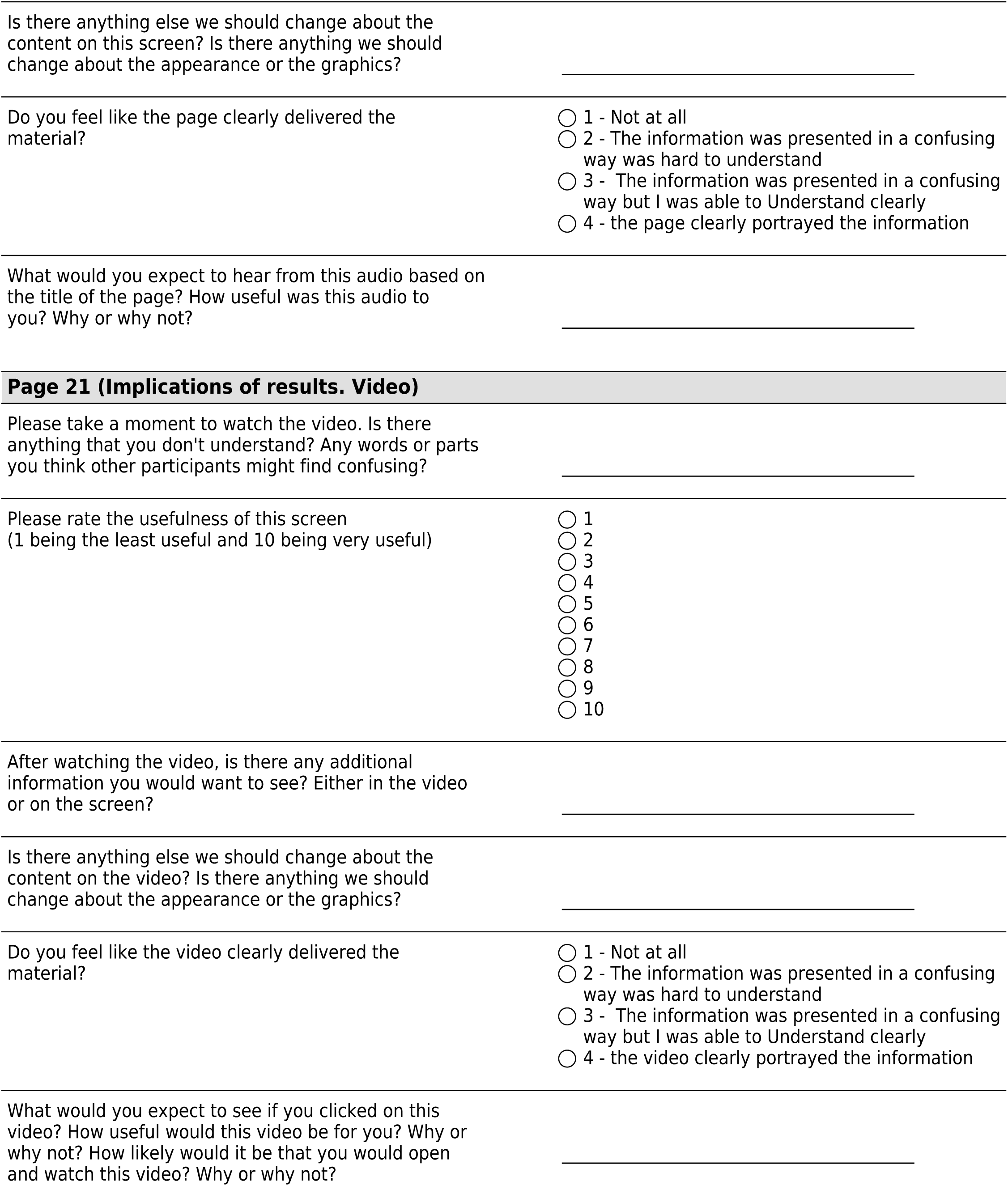
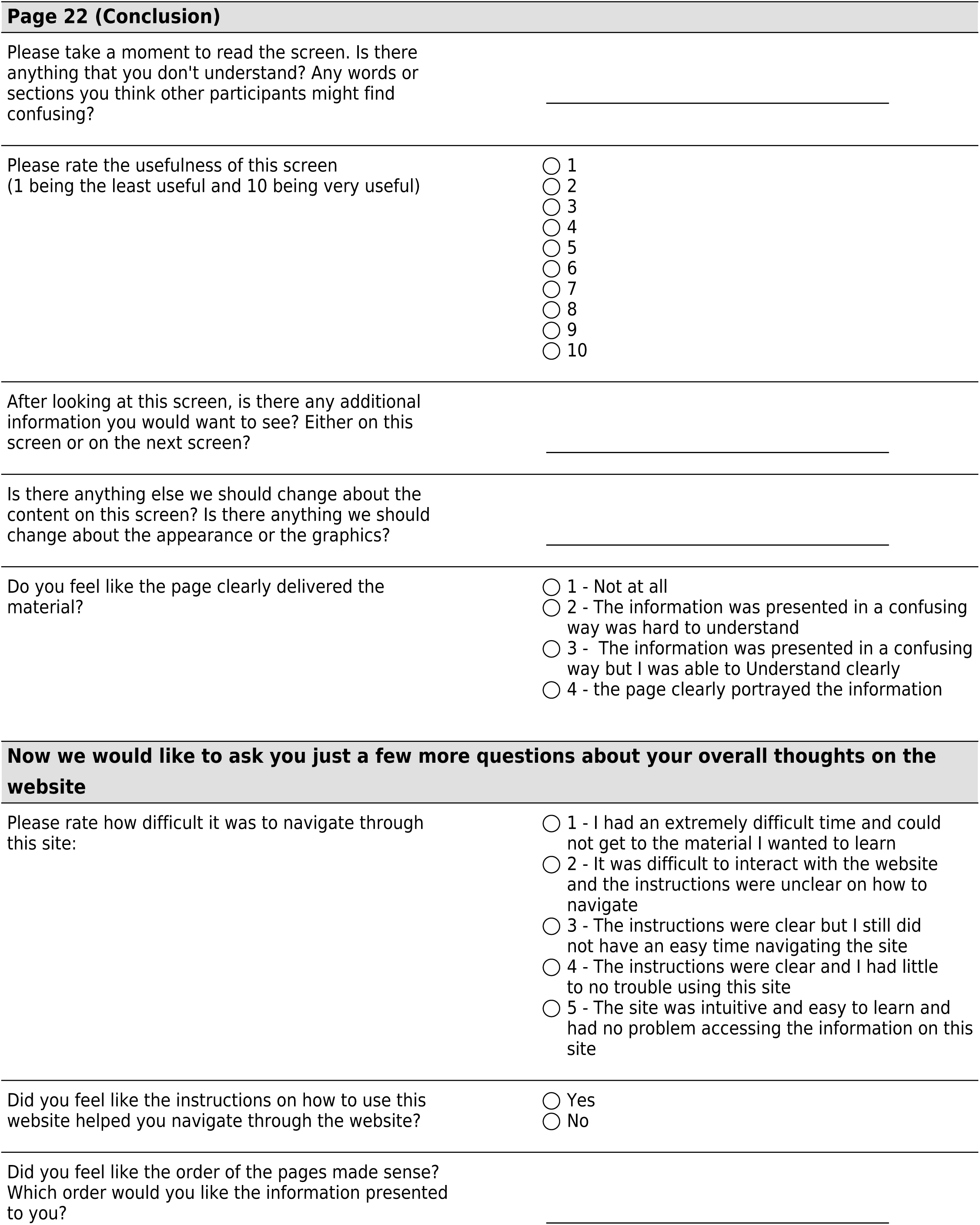
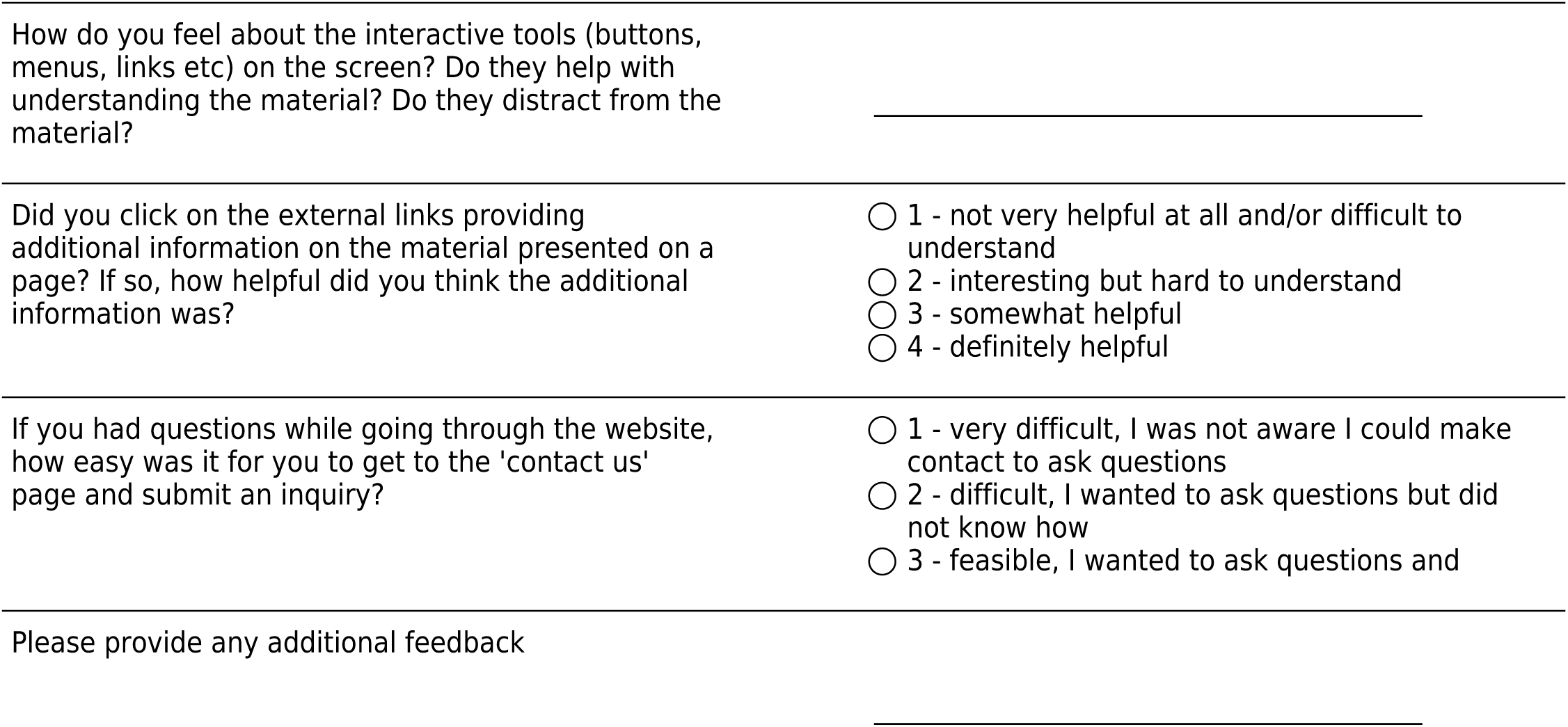

## Usability Questions

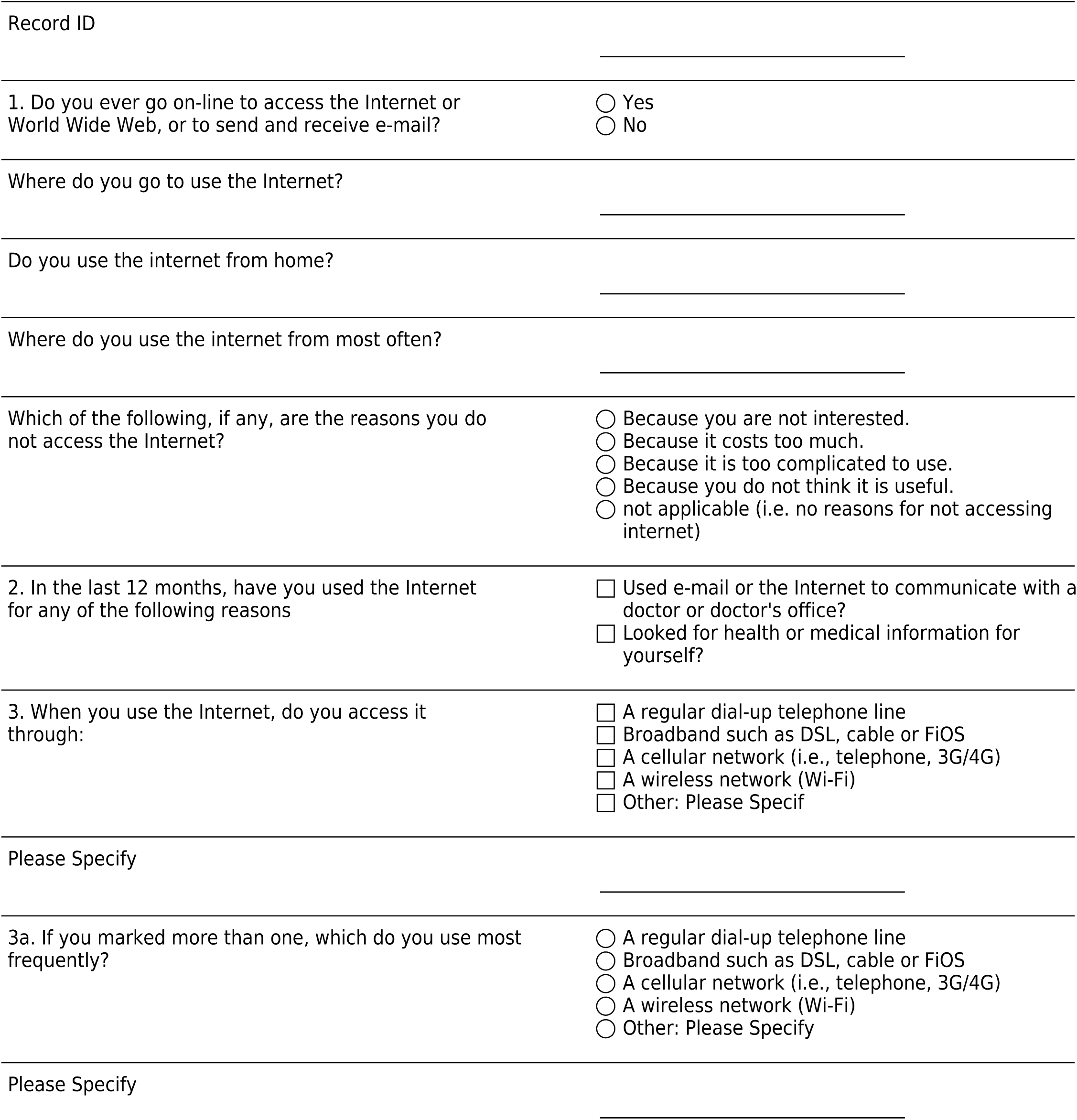
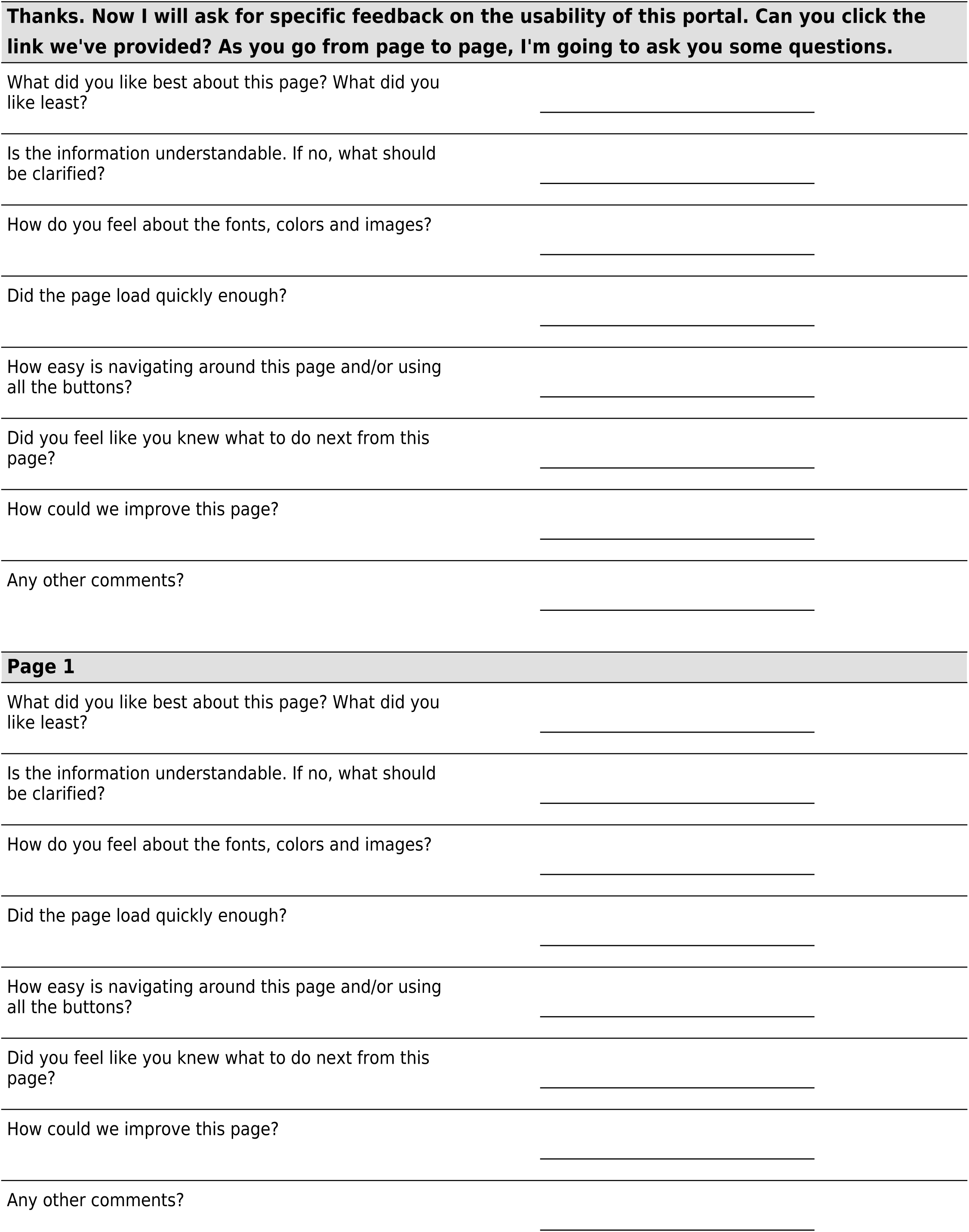
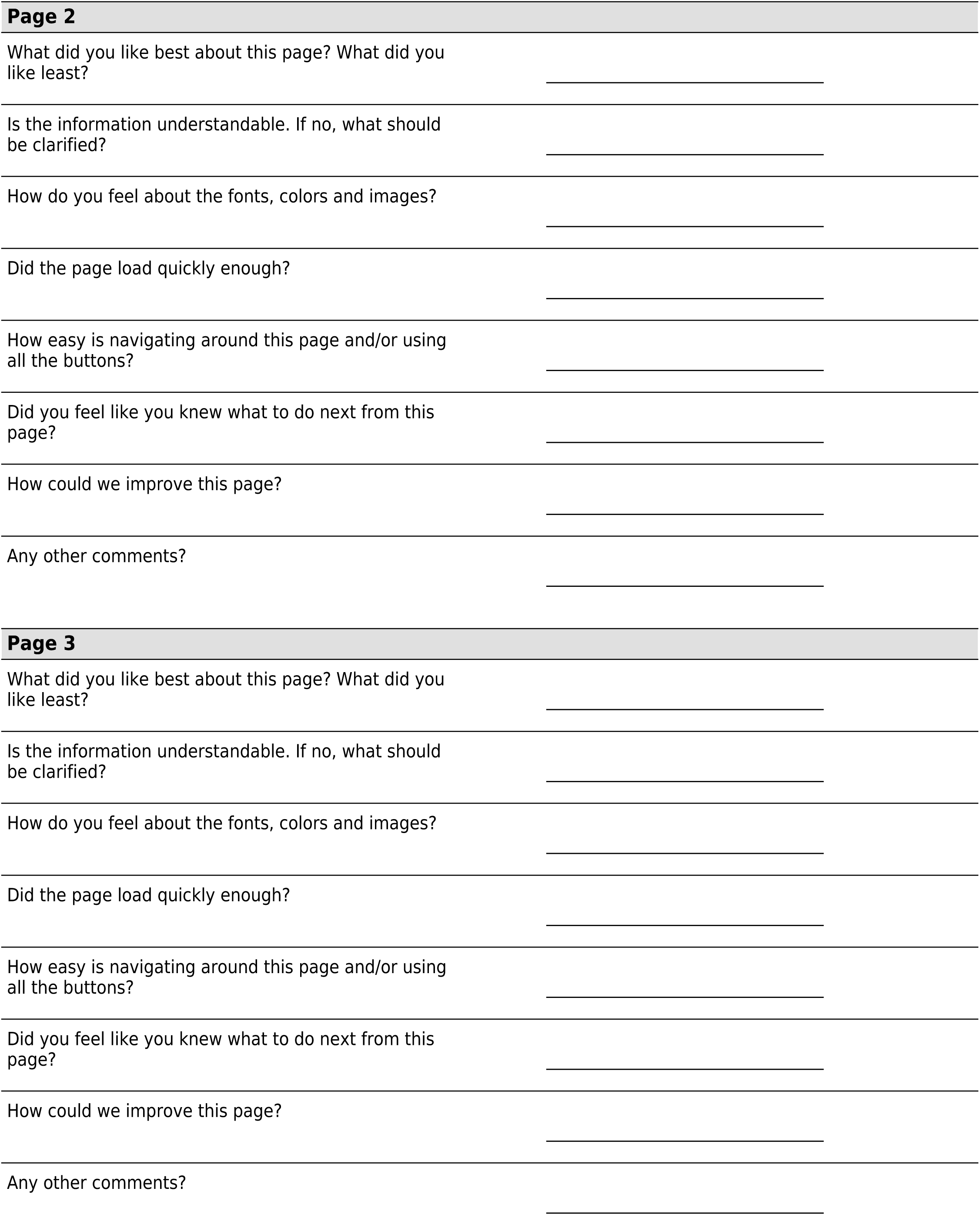
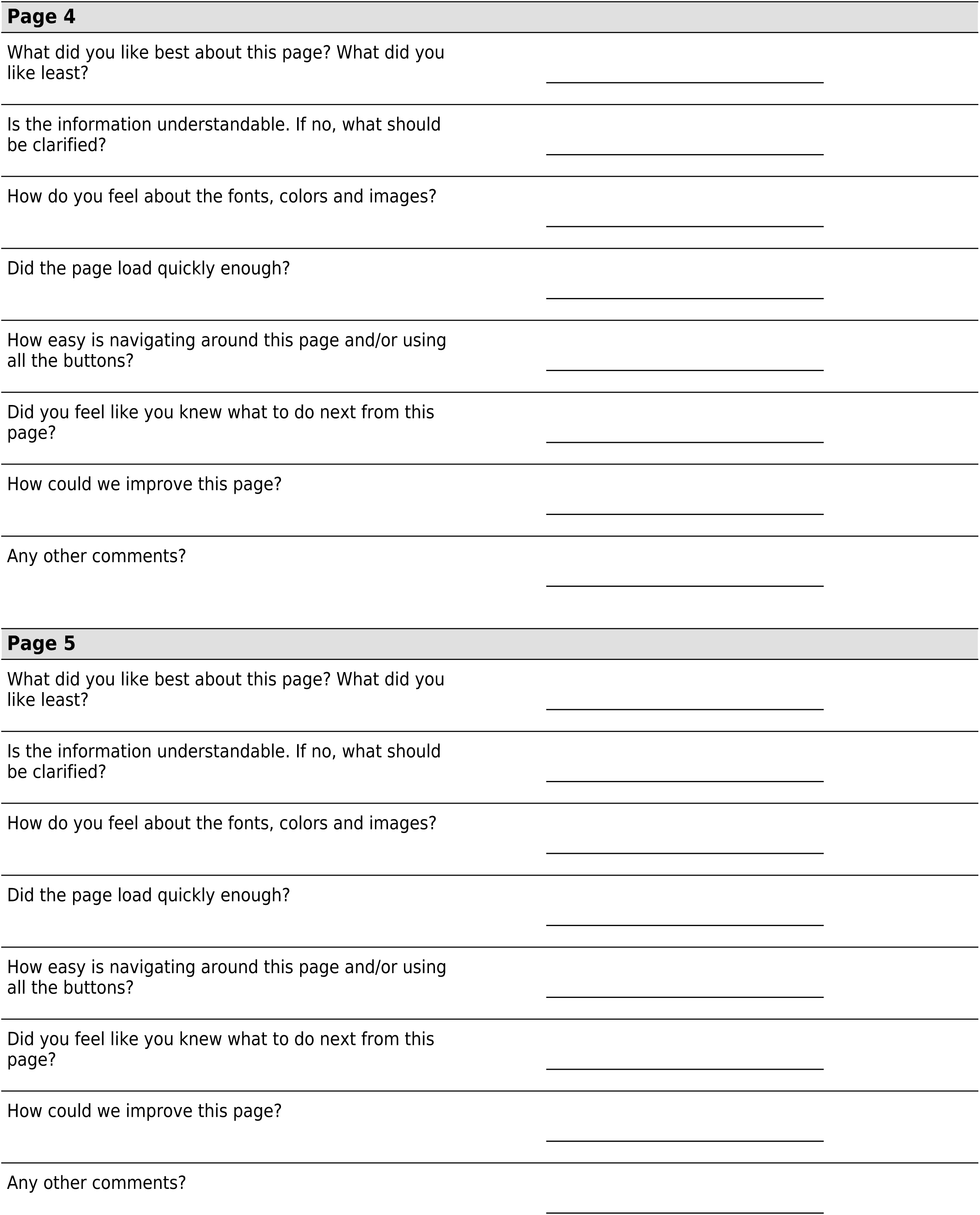
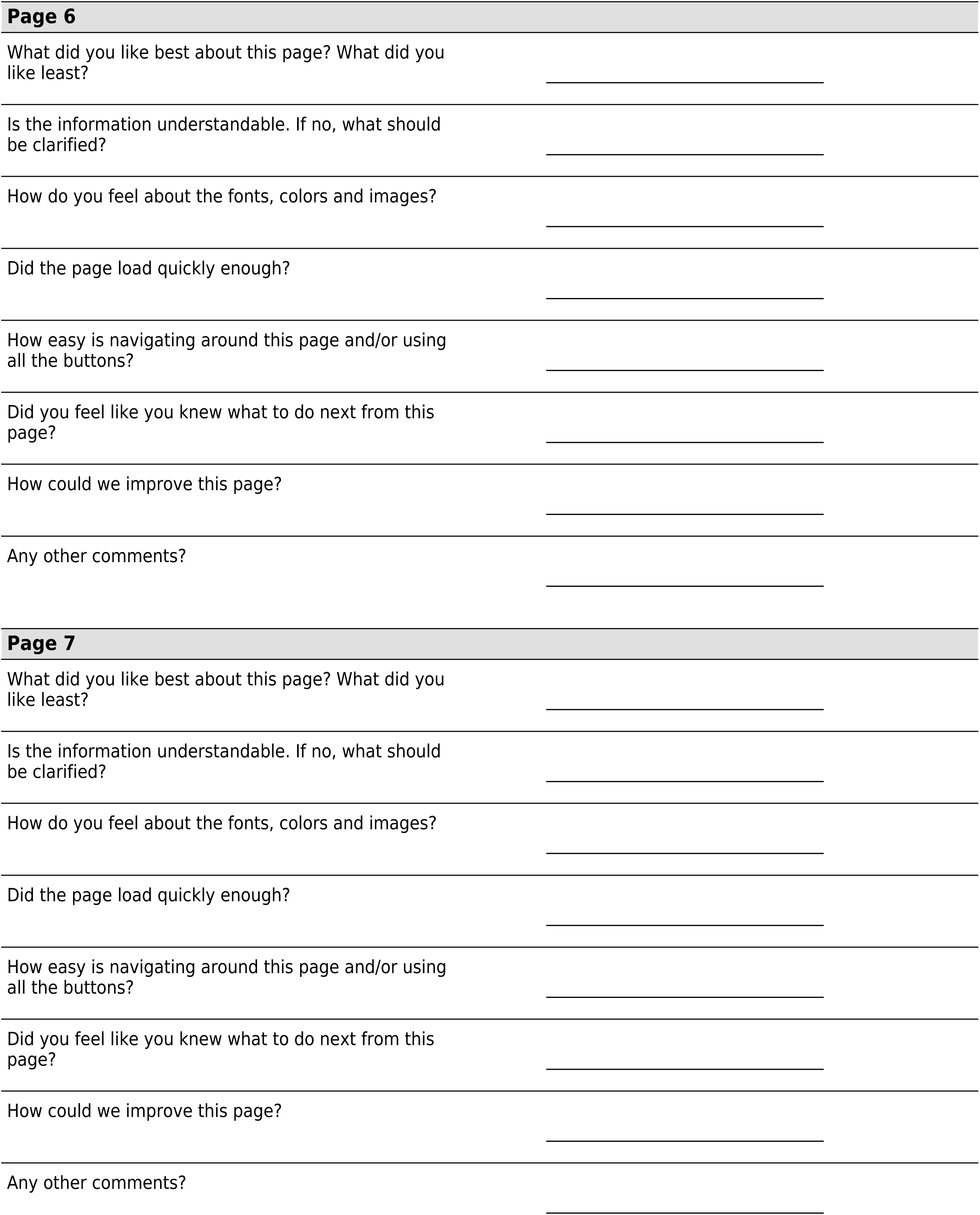
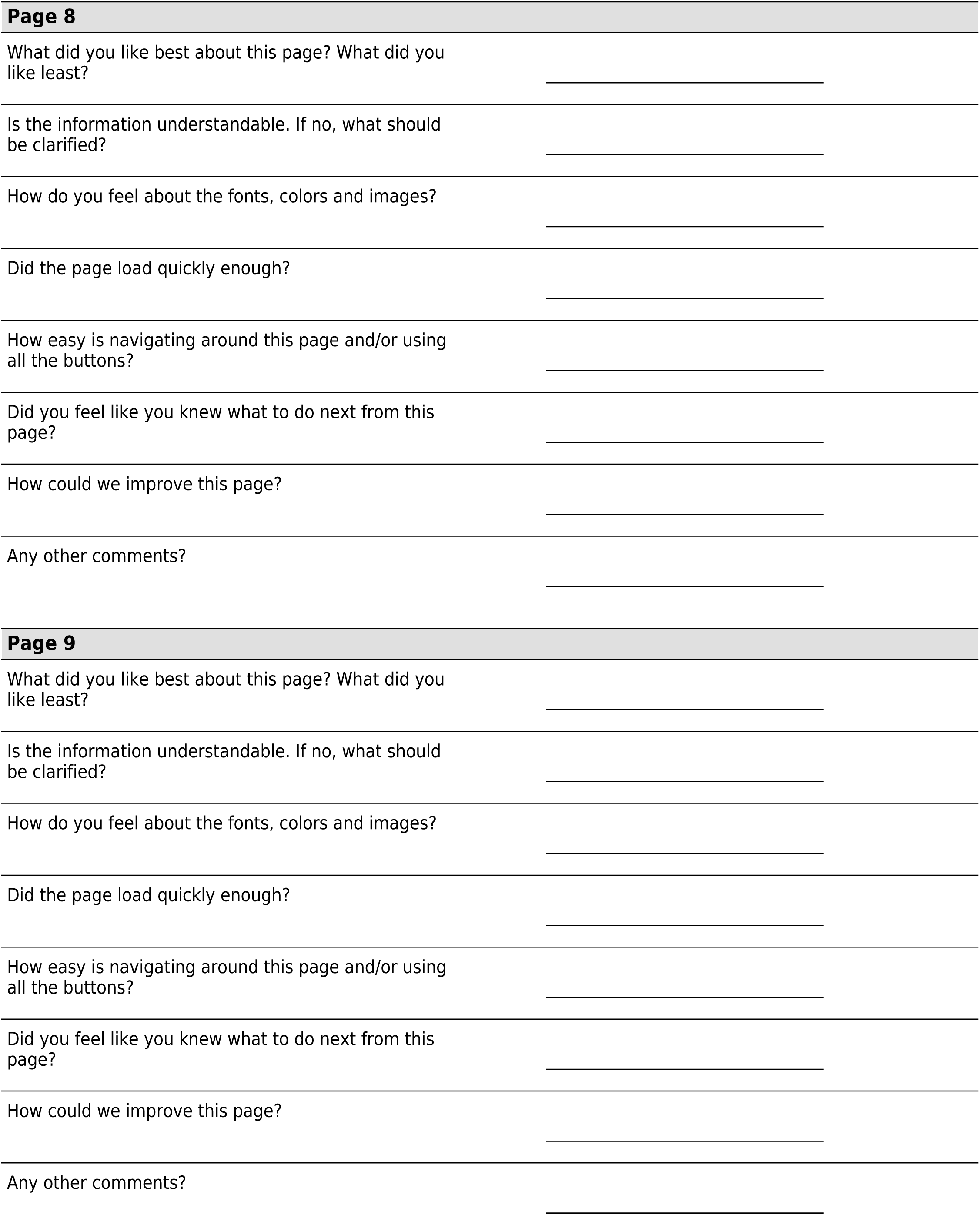
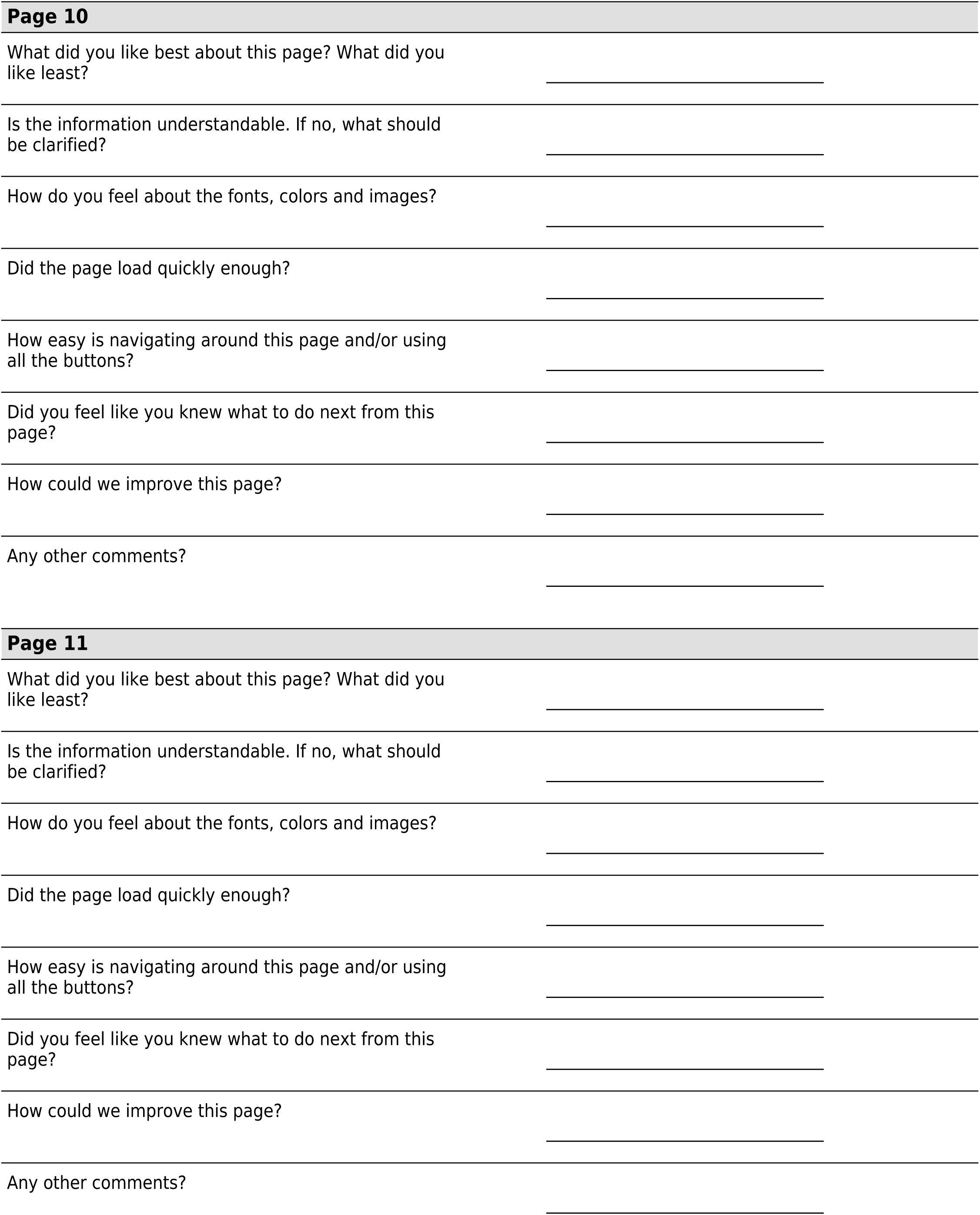
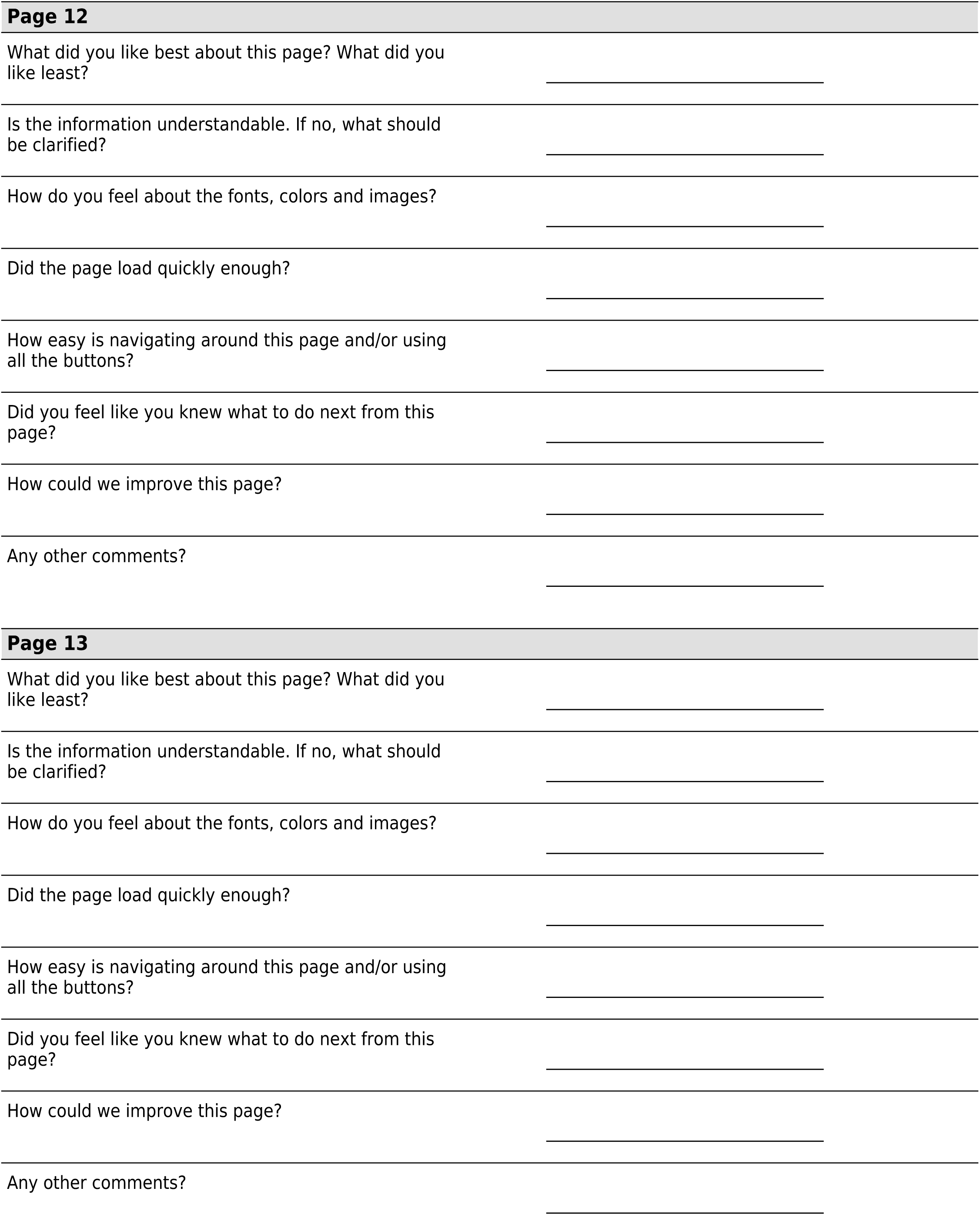
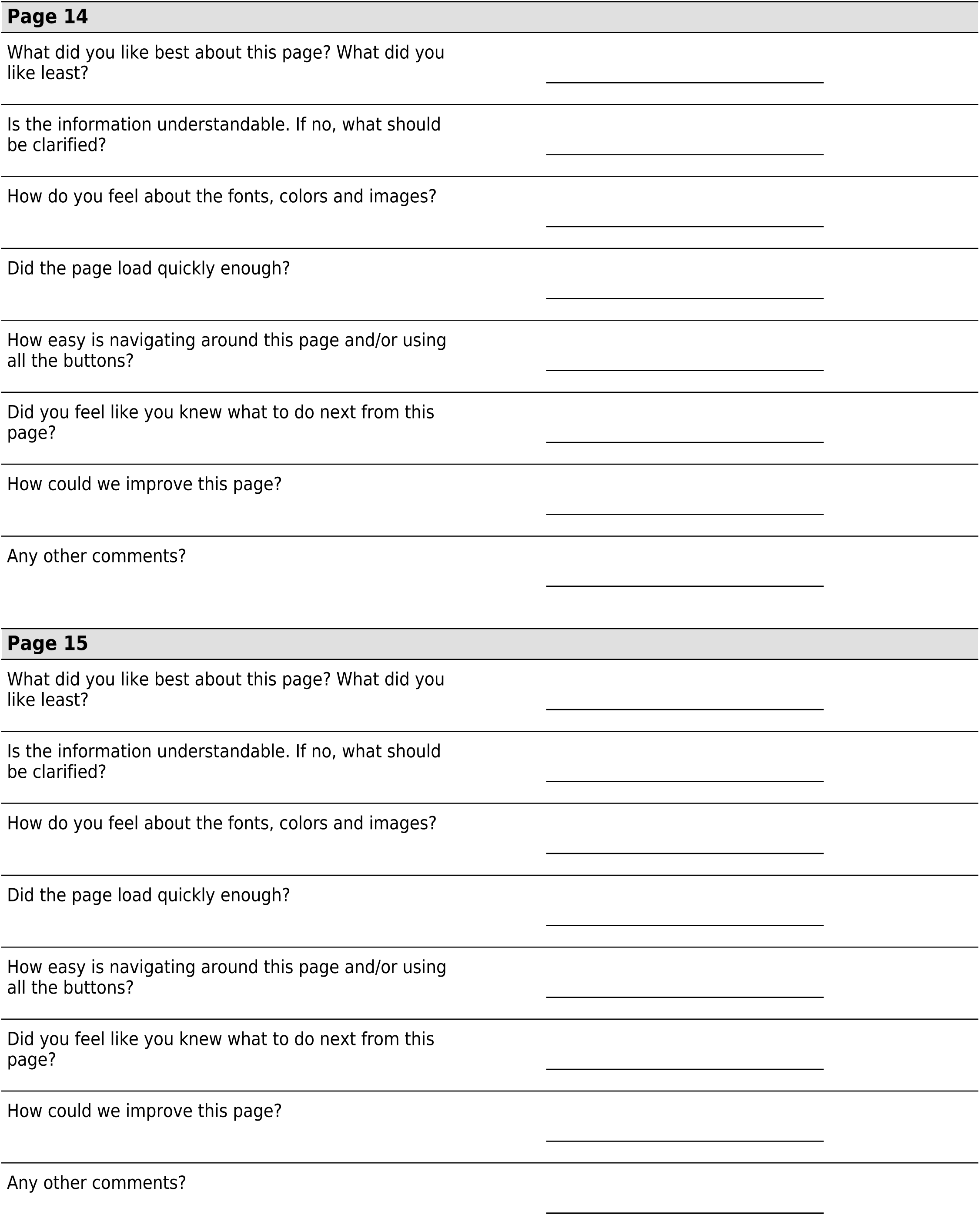
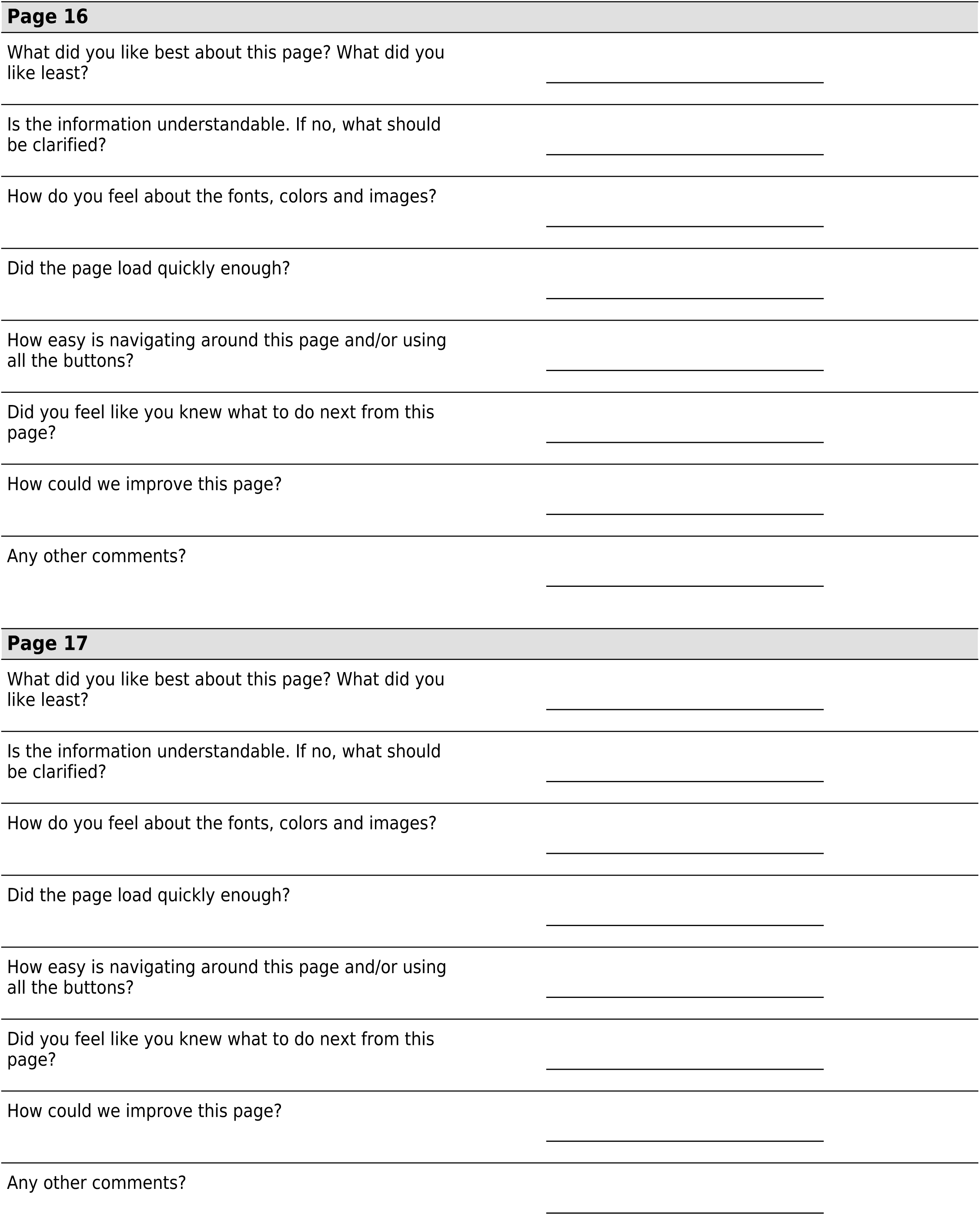
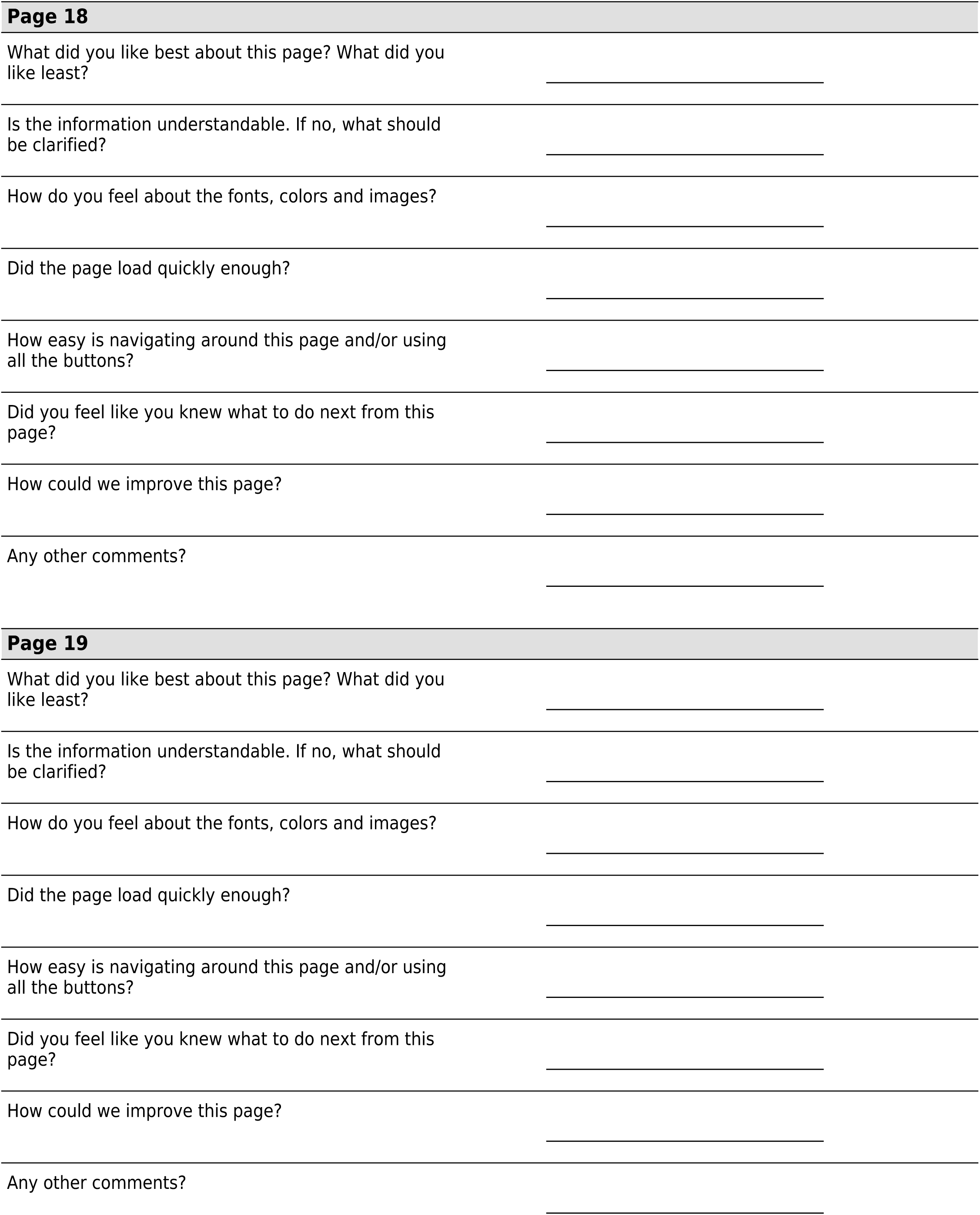

## Notes

### Competing Interest Statement

The authors have declared no competing interest.

### Funding Statement

This study was funded by The University of Pennsylvania

### Author Declarations

IRB of The University of Pennsylvania gave ethical approval for this work

