## Supplemental Materials for "User and Usability Testing of a web-based *GBA* and *LRRK2* genetics education tool"

by **Han et al.**

**Includes:**

Supplemental Table 1.

Supplemental Table 2.

Supplemental Table 3.

Supplemental Table 4.

Supplemental Questionnaire 1.

Supplemental Questionnaire 2.

**Supplemental Table 1.** Internet use reported by movement disorders specialists (N=11) and people with Parkinson's disease (N=13) in the content review (Phase 1). Available data presented as precent of total cohort.

| | Movement Disorders Specialist | Person with Parkinson's Disease |
|---|---|---|
| **Internet Use** | | |
| >12 hours per week | 9 (69%) | 7 (54%) |
| 10-12 hours/week | 1 (9%) | 3 (23%) |
| 7-9 hours/week | 1 (9%) | 3 (23%) |
| **What device do you use for internet access?** (more than once response allowed) | | |
| Smartphone | 11 (100%) | 13 (100%) |
| Desktop or Laptop | 11 (100%) | 11 (85%) |
| Tablet | 6 (55%) | 6 (46%) |
| **Type of internet access** (more than once response allowed) | | |
| Dial-up | 0 (0%) | 0 (0%) |
| DSL or Broadband (wired or wi-fi) | 11 (100%) | 13 (100%) |
| Cellular network | 7 (64%) | 6 (46%) |
| **Comfort with internet use** | | |
| Confident/no need for help | 8 (73%) | 5 (35%) |
| Comfortable/rare assistance | 3 (27%) | 4 (31%) |
| Need some assistance | 0 (0%) | 3 (23%) |
| Uncomfortable/needs assistance | 0 (0%) | 1 (8%) |

**Supplemental Table 2.** Average usefulness scale for each page reported by movement disorders specialists (N=11) and people with Parkinson's disease (N=13) in the content review (Phase 1). Usefulness is reported from 1-10 with 1 being the lowest and 10 being the highest. PD = Parkinson's disease. GT = Genetic Testing. VUS = Variant of uncertain significance.

| Page | Movement Disorders Specialist | Person with Parkinson's Disease |
|---|---|---|
| 1. Title | 8.36 | 6.85 |
| 2 How to use the website | 9.27 | 8.77 |
| 3. Introduction | 9.45 | 8.62 |
| 4. Content Summary | 9.70 | 9.15 |
| 5. What is PD? | 8.55 | 9.08 |
| 6. PD Signs and Symptoms | 8.18 | 8.00 |
| 7. Causes of PD | 8.64 | 8.62 |
| 8. PD Treatments | 7.20 | 8.62 |
| 9. Inheritance | 8.36 | 8.77 |
| 10. When to consider GT | 9.20 | 8.23 |
| 11. Additional info about GT | 7.20 | 9.17 |
| 12. Genetics Introduction | 8.73 | 9.38 |
| 13. Types of GT | 9.30 | 8.31 |
| 14 Risks and Benefits of GT | 9.50 | 8.62 |
| 15. GT Process | 9.30 | 7.62 |
| 16. Role of Genetics in PD | 9.00 | 8.85 |
| 17. GBA | 9.11 | 8.54 |
| 18. LRRK2 | 9.11 | 8.82 |
| 19. Other PD Genes | 8.89 | 7.82 |
| 20 VUS | 8.70 | 8.54 |
| 21 How do I get my results | 9.22 | 7.85 |
| 22. Implications/limitations | 9.10 | 7.46 |
| 23. Conclusion | 9.44 | 9.73 |
| **Average** | 8.85 | 8.49 |
| **Standard Deviation** | 0.64 | 0.65 |

**Supplemental Table 3.** Summary feedback reported by movement disorders specialists (N=11) and people with Parkinson's disease (N=13) in the content review (Phase 1). Available data presented as percent of total cohort.

| | Movement Disorders Specialist | Person with Parkinson's Disease |
|---|---|---|
| **Website navigation was:** | | |
| 'Intuitive and easy to learn and had no problem accessing the information on this site' | 6 (55%) | 10 (77%) |
| 'the instructions were clear, and I had little to no trouble using this site' | 4 (36%) | 3 (23%) |
| **The information found on external links was:** | | |
| Definitely helpful | 6 (55%) | 7 (54%) |
| Somewhat helpful | 0 (0%) | 2 (15%) |
| Interesting but hard to understand | 1 (9%) | 0 (15%) |
| Not very helpful and/or difficult to understand | 2 (18%) | 0 (15%) |
| **Access to the 'contact us' page was:** | | |
| Feasible | 9 (82%) | 10 (77%) |
| Difficult | 0 (0%) | 2 (15%) |
| I was not aware I could make contact to ask questions | 1 (9%) | 1 (8%) |

**Supplemental Table 4**. Final remarks from movement disorders specialists (N=11) and people with Parkinson's disease (N=13) in the content review (Phase 1).

| Movement Disorders Specialist Feedback | | | | |
|---|---|---|---|---|
| Page | **Did you feel like the order of the pages made sense? Which order would you like the information presented to you?** | **How do you feel about the interactive tools? Did they help with understanding the material? Do they distract from the material?** | **Did you click on the external links providing additional information on the material presented on a page? If so, how helpful did you think the additional information was? (Scale of 1-4)** | **Please provide any additional feedback** |
| 1 | See above, for the most part yes. | They were helpful | 4 - definitely helpful | It's great! |
| 2 | look above | helpful because it breaks up the info | 4 - definitely helpful | No |
| 3 | order made sense | overall was helpful but needs tweaks. | 1 - not very helpful at all and/or difficult to understand | nope |
| 4 | See above. | helpful. | 4 - definitely helpful | Have a progress bar. have direction on first page directing the order of actions, listen then read or vice versa. |
| 5 | Yes | Helpful | 4 - definitely helpful | for pics have 1 AA, 1 Hispanic, 1 SE Asian, 1 East Asian. may want to ask whether someone had to help them navigate through the site and that its okay to have someone there helping through the site |
| 6 | Just one page, look above | more helpful, can be more interactive | | talk a little more about how to get results back. maybe more obvious method of contact mentions the more personal side of why people choose or don't choose to get genetic testing (testimonials) |
| 7 | They made sense | helpful | 2 - interesting but hard to understand | main point of this website is to focus on genetics and work backwards from the main point to streamline the website. |
| 8 | Yes. autosomal and dominant and recessive should be | helpful | 4 - definitely helpful | maybe make contact us more obvious |

| | | | | |
|---|---|---|---|---|
| | moved up earlier, the nature of the testing may also need to move up earlier | | | |
| 9 | Yes | at times they were good but need to be synced | 4 - definitely helpful | Make sure purpose is clear, is it for PD genetic testing in general or just for MIND |
| 10 | | | | |
| 11 | Yes | Helpful | 1 - not very helpful at all and/or difficult to understand | make sure we aren't overwhelming patients, let them know how long it’s going to take |
| People with Parkinson’s Disease Feedback | | | | |
| 1 | Yes | distracting if the text didn't match the audio or if it was hard to read | 3 - somewhat helpful | No |
| 2 | Yes | Helped | | No |
| 3 | order made sense | helpful | 4 - definitely helpful | have audio more obvious on the page. |
| 4 | Yes | a little bit of both | | be more selective on information. focus more on privacy |
| 5 | Yes | helpful | | make the purpose of this study clearer |
| 6 | Yes | helpful | 4 - definitely helpful | No |
| 7 | Yes | Helpful tools | 4 - definitely helpful | Make the play button for audio and video bigger |
| 8 | Made sense | helpful | | make the website flow more smoothly. |
| 9 | It made sense | Yes | 3 - somewhat helpful | Shorter. May need to cut some out. |
| 10 | look above, add a summary | they were fine | 4 - definitely helpful | No |
| 11 | made sense | helpful but should be more obvious when to scroll to next piece of info | 4 - definitely helpful | No |
| 12 | Look above | helpful | 4 - definitely helpful | No |
| 13 | Order was fine | more distracting | 4 - definitely helpful | make it more visual appealing |
